# DNA methylome-wide association study of genetic risk for depression implicates antigen processing and immune responses

**DOI:** 10.1101/2021.06.30.21259731

**Authors:** Xueyi Shen, Doretta Caramaschi, Mark J Adams, Rosie M Walker, Josine L Min, Alex Kwong, Gibran Hemani, Genetics of DNA Methylation Consortium, Miruna C Barbu, Heather C Whalley, Sarah E Harris, Ian J Deary, Stewart W Morris, Simon R Cox, Caroline L Relton, Riccardo E Marioni, Kathryn L Evans, Andrew M McIntosh

## Abstract

**Background:** Depression is a disabling and highly prevalent condition where genetic and epigenetic differences, such as DNA methylation (DNAm), contribute to prediction of disease liability.

**Method:** We investigated the association between polygenic risk scores (PRS) for depression and DNAm by conducting a methylome-wide association study (MWAS) in Generation Scotland (N=8,898, mean age=49.8 years) with replication in the Lothian Birth Cohorts of 1921 and 1936 and adults in Avon Longitudinal Study of Parents and Children (ALSPAC) (N_combined_=2,049, mean age=79.1, 69.6 and 47.2 years, respectively). We also conducted a replication MWAS in the ALSPAC children (N=423, mean age=17.1 years).

**Result:** Wide-spread associations were found between PRS constructed using genetic risk variants for depression and DNAm in cytosine-guanine dinucleotide (CpG) probes that localised to genes involved in immune responses and neural development (N_CpG_=599, p_Bonferroni_<0.05, p<6.5×10^−8^). The effect sizes for the significant associations were highly correlated between the discovery and replication samples in adults (r=0.83) and in adolescents (r=0.76). Additional analysis on the methylome-wide associations was conducted for each lead genetic risk variant. Over 40% of the independent genetic risk variants showed associations with CpG probe DNAm located in both the same (*cis*) and distal probes (*trans*) to the genetic loci (p_Bonferroni_<0.045). Subsequent Mendelian randomisation analysis showed that DNAm and depression are mutually causal (p_FDR_<0.039), and there is a greater number of causal effects found from DNAm to depression (DNAm to depression: p_FDR_ ranged from 0.045 to 2.06×10^−120^; depression to DNAm: p_FDR_ ranged from 0.046 to 2.1×10^−23^).

**Conclusion:** Polygenic risk scores for depression, especially those constructed from genome-wide significant genetic risk variants, showed epigenome-wide methylation association differences in the methylome associated with immune responses and brain development. We also found evidence from Mendelian randomisation evidence that DNAm may be causal to depression, as well as a causal consequence of depression.

## Introduction

Depression is a highly prevalent condition and a leading cause of global disability^1^, for which the underlying biological mechanisms are unclear. Genetic factors account for a substantial proportion of differences in liability to depression, which has a twin-based heritability of approximately 37% and with common genetic variants capturing around 6-10% of phenotypic variance^2,3^. Recent genome-wide association studies (GWAS) have identified specific genetic risk variants for depression that implicate regional brain alterations^4,5^. Polygenic risk scores (PRS) derived from the results of GWAS studies, have been widely used to estimate additive genetic risk^6^. PRS provides a means to identify traits that share their genetic architecture with depression, which may help to prioritise factors of biological and mechanistic relevance for the disorder^7^.

DNA methylation (DNAm) at cytosine-guanine dinucleotides (CpG) sites is one of the most studied epigenetic markers and there is growing evidence of its role in understanding depression^8^. DNAm risk scores have been developed from the results of DNA methylome-wide association studies (MWAS)^8^. These can be used to predict prevalent depression in independent samples, and chronic depression that requires long-term treatment^9^. DNAm is influenced by both genetic and environmental factors ^8,10^ and, in blood tissue, it has a mean heritability of 19% across the epigenome^11^ with ∼7% of its variance captured by common genetic variants^12^. For the highly heritable DNA methylation probes, genetic effects are consistent across tissues^13^ and developmental stages^12^. Genetic risk variants for diseases, such as schizophrenia, have been found enriched in DNA methylation variation^14–16^. Associations between genetic risk and epigenetic changes can enrich our understanding of the functional composition of genetic risk loci, and thus inform the mechanisms that lead to the onset of depression^17,18^. However, systematic examination of the molecular genetic associations between genetic risk of depression and DNAm has not, to the best of our knowledge, been conducted.

In the present study, we aim to investigate the association between PRS for depression and genome-wide DNA methylation. Methylome-wide association studies (MWAS) were conducted on four cohorts: Generation Scotland: Scottish Family Health Study (GS, discovery sample, N=8,898), the Lothian Birth Cohort (LBC1921), the Lothian Birth Cohort 1936 (LBC1936), Avon Longitudinal Study of Parents and Children (ALSPAC) adults (adult replication sample, combined N=2,049) and ALSPAC children for replication (adolescent replication sample, N=423). Mendelian randomisation was used to test for causal associations between DNAm and depression using data from the Genetics of DNA Methylation Consortium (GoDMC) (N=25,561) and GS.

## Methods

### Sample descriptions

#### GS

Generation Scotland (GS) is a family-based population cohort with over 24,000 participants^19,20^ set-up to identify the causes of common complex disorders, such as depression. DNA methylation data and genetic data were both collected, processed and quality-checked for 8,898 people (mean age=49.8 years, SD of age=13.7 years, 40.90% were men) in two sets. Sample sizes for set 1 and set 2 were 4757 (mean age=48.5 years, SD of age=14.0 years, 38.5% were men) and 4141 (mean age=51.4 years, SD of age=13.2 years, 43.66% were men), respectively. Written informed consent was obtained for all participants. The study was approved by the NHS Tayside Research Ethics committee (05/s1401/89).

#### Lothian Birth Cohorts (LBC) 1921 and 1936

Participants from LBC 1921 and LBC 1936^21,22^ were born in 1921 and 1936. Almost all lived in the Edinburgh and surrounding Lothian area when recruited. They are a mostly healthy, community-dwelling sample of men and women. The sample used in the current analysis included 1,330 participants from both cohorts combined with genetic and DNA methylation data (LBC 1921: mean age=79.1 years, SD of age=0.6, 39.7% were men; LBC 1936: mean age=69.6 years, SD of age=0.8, 50.6% were men; all participants were unrelated). Written informed consent was obtained from all participants. Ethics permission for LBC1921 was obtained from the Lothian Research Ethics Committee (LREC/1998/4/183). Ethics permission for LBC1936 was obtained from the Multi-Centre Research Ethics Committee for Scotland (MREC/01/0/56) and the Lothian Research Ethics Committee (LREC/2003/2/29)^23,24^.

#### Avon Longitudinal Study of Parents and Children (ALSPAC)

The Avon Longitudinal Study of Parents and Children (ALSPAC) is an ongoing longitudinal population-based study that recruited pregnant women residing in Avon (South-West of England) with expected delivery dates between 1st April 1991 and 31st December 1992^25,26^. The cohort consists of 13,761 mothers and their partners, and their 14,901 children, now young adults^27^. The study website contains details of all the data that is available through a fully searchable data dictionary and variable search tool (http://www.bristol.ac.uk/alspac/researchers/our-data/). Ethical approval for the study was obtained from the ALSPAC Ethics and Law Committee and the Local Research Ethics Committees. A subsample of 719 unrelated mothers with DNA methylation (DNAm) data (mean age=47.2 years, SD of age=4.6) were included in the replication study^28^. Supplementary analyses were also conducted on a younger subsample with DNAm consisting of 423 young people (mean age=17.1 years, SD of age=1.1 and 41% were boys). Details of the selection of participants for these subsamples are in the study by Relton et al^28^. Consent for biological samples has been collected in accordance with the Human Tissue Act (2004).

### Genotyping and imputation

Detailed information on the quality control and genotyping methods for GS^19^, LBC1921, LBC1936^29^ and ALSPAC^30^ have been previously published and is described briefly below. Analyses were conducted on European participants.

#### GS

Each sample was genotyped using the IlluminaHumanOmniExpressExome-8v1.0 BeadChip (48.8%) or Illumina HumanOmniExpressExome-8 v1.2 BeadChip (51.2%) with Infinum chemistry^31^. Quality control included removing participants with genotyping call rate<98%, SNP removal of those with a minor allele frequency (MAF)<1%, call rate<98%, Hardy-Weinberg equilibrium (HWE) p-value<5×10^−6^. Imputation was performed using the Sanger Imputation server with the Haplotype Reference Consortium reference panel (HRC.r1-1). SNPs with INFO score<0.8 were removed from analysis.

#### LBC1921 and LBC1936

Genotyping was performed using the Illumina610-Quadv1 chip (Illumina, Inc., San Diego, CA, USA). Participants were excluded with a call rate<95%. SNPs were removed if MAP<5%, call rate<98%, HWE p-value<0.001. Imputation and quality control based on INFO score were the same as GS.

#### ALSPAC

Genotyping arrays used were the Illumina Human660W-quad chip for mothers and Illumina HumanHap550-quad chip for children. SNPs with missingness>0.05, HWE p-value<1×10^−6^, MAF<0.01 were excluded. The above quality control steps were conducted on the entire genotyped sample. Imputation and quality control based on INFO score were consistent with similar procedures used in GS.

### Polygenic profiling

Polygenic risk scores (PRS) of depression were calculated using PRSice-2^32^ for GS, LBC1921, LBC1936 and ALSPAC separately, using the summary statistics of a genome-wide meta-analysis of depression by Howard *et al*.^33^ excluding individuals from GS previously included in that GWAS meta-analysis. Nine p-value cut-offs were used for thresholding SNPs in the summary statistics (pT): 1, 0.5, 0.1, 0.05, 0.01, 1×10^−3^, 1×10^−4^, 1×10^−5^ and 5×10^−8^ for clumping and thresholding. Each set of SNPs was used to generate a depression-PRS in GS. A separate PRS was generated using the lead genetic risk variants or their closest proxies (in LD r^2^>0.1) reported in the GWAS by Howard *et al*.^33^ for supplementary analysis. Details of the PRS profiling procedures can be found elsewhere^33^ (also see Supplementary Information and Supplementary Table 1-2).

Subsequently, using the lead risk variants reported by Howard et al^33^, we tested for individual SNP-CpG associations in GS. Lead risk variants were selected by extracting the most significant proxy SNPs (p<5×10^−8^) in linkage disequilibrium (LD r^2^>0.01) with the lead variants reported in the Howard *et al.* study^4^. A total of 96 SNPs were available and thus selected as leading risk variants for further analysis.

### DNA methylation data

#### GS

Genome-wide DNA methylation data was obtained from whole blood samples using the Illumina Infinium Methylation EPIC array (https://emea.support.illumina.com/array/array_kits/infinium-methylationepic-beadchip-kit.html). Data processing was performed separately for each set. Quality control (QC) and normalisation were conducted using R packages ‘ShinyMethyl’ (version 1.28.0)^34^, ‘watermelon’ (version 1.36.0)^35^ and ‘meffil’ (version 1.1.1)^36^. Details of the protocol are described elsewhere^37^. In summary, quality control procedures removed probes if there was an outlying log median methylated signal intensity against unmethylated signal for each array, or a bead count<3 in ≥5 % of the total probe sample, or a detection p-value>0.05 for set 1 and p-value>0.01 for set 2 in ≥0.5% of the total sample in each respective set. Cross-hybridising probes that map to genetic variants at MAF>0.05 and polymorphic probes were removed^38^. Samples were excluded if sex prediction from methylation data was inconsistent with self-reported data, or a detection p-value >0.05 for set 1 and p-value>0.01 for set 2 found in >1% of the overall probes for each set respectively. The data was then normalised using the ‘dasen’ method from the ‘waterRmelon’ R package (version 1.36.0).

The raw intensities were then transformed into M-values by log-transforming the proportional methylation intensity^39^. The M-values were corrected using a linear-mixed model, controlling for relatedness using the GCTA-estimated genetic relationship matrix^40^ for set 1. This step was omitted for set 2 as all participants were unrelated within the set and to set 1. The residualised M-values for over 750,000 autosomal CpG probes were then used for further analysis.

#### LBC1921 and LBC1936

Genome-wide DNA methylation data was obtained from blood sample using the HumanMethylation450K array (https://emea.illumina.com/content/dam/illumina-marketing/documents/products/datasheets/datasheet_humanmethylation450.pdf)^41,42^. Quality control and normalisation were performed using the ‘minfi’ R package (version 1.38.0)^41^. Probes with low call rate (<95%), outlying M-values (>3 SD from mean) or identified as cross-hybridising and polymorphic were removed^43^. Participants with insufficient cell count information were excluded from analysis.

All participants in LBC1921 and LBC1936 with methylation data were unrelated. M-value transformation were conducted consistently with the GS sample. Data for over 450,000 CpG probes were retained for further analysis.

#### ALSPAC

Illumina Infinium HumanMethylation450 Beadchip arrays were used for measuring genome-wide DNA methylation data from peripheral blood sample. The R package ‘meffil’ (version 1.1.1) was used for pre-processing, quality control and normalisation as previously reported^44^. Further removal of probes was conducted based on background detection (p>0.05) and if they reached beyond the 3 times inter-quantile range from 25% and 75% or identified as cross-hybridising or polymorphic^43^. Related (IBD>0.1) participants were not included in the analyses^30^.

M-value transformation was conducted and over 468,000 CpG probes remained for analysis.

### Statistical models for methylome-wide association analysis (MWAS)

A discovery MWAS was initially conducted in GS. Two separate analyses were conducted on sets 1 and 2, and the final summary statistics were obtained by meta-analysing the two sets of results using METAL (version released in 2011)^45^. We used the default analysis scheme without genomic control correction (genomic inflation factors reported in the Supplementary Information). P-values for the meta-analysis were obtained from a fixed-effect inverse-variance model. A replication analysis on adults was then conducted on the total sample from LBC1921, LBC1936 and ALSPAC adults. Replication MWAS was first conducted separately for each cohort and then meta-analysed using the same parameters for the discovery analysis. Finally, an additional replication MWAS was conducted on ALSPAC children. All analyses were conducted using R (version 3.5.1) under Linux environment.

Linear regression was used to test the associations between depression-PRS and for each CpG using the R package ‘limma’^46^ (version 3.48.0) for GS, LBC1921 and LBC1936. The ‘lmFit’ function was first implemented to test the association for each CpG. The inference statistics of each linear model was then adjusted using the ‘eBayes’ function, by which an empirical Bayes method was used to adjust for gene-wise variance using a shrinkage factor. Moderated t-statistics and p-values were produced by this step. In ALSPAC, the analyses were conducted using the ‘meffil’ (version 1.1.1) R package, using the ‘sva’ option^44^.

Self-reported smoking status, smoking pack years, DNAm-estimated white-blood cell proportions (CD8+T, CD4+T, natural killer cells, B cells and granulocytes)^37^, batch, the first 20 principal component derived from the M-values, age and sex were included as covariates for the discovery methylome-wide association analysis (see Supplementary Information and Supplementary Table 3 for details). Where possible, the same covariates were used in the replication analyses, although only smoking status (and not pack years) were available in LBC 1921, LBC 1936 and ALSPAC. Details for all the covariates included in the replication analysis can be found in the Supplementary Information. MWAS were conducted for the nine depression-PRS scores separately. P-values were Bonferroni-corrected (p-value threshold = 6.5×10^−8^ for EPIC array used in GS, 1.1×10^−7^ for 450k array used for replication analysis in LBC1921, LBC1936 and ALSPAC adults, and 1.1×10^−7^ for replication in ALSPAC children). Standardised regression coefficients are reported as effect sizes. For the significant CpG probes, gene symbol annotation and UCSC classification of CpG Island positions were acquired from the ‘UCSC_RefGene_Name’ and ‘Relation_to_Island’ columns, respectively, from the annotation object generated by the ‘IlluminaHumanMethylationEPICanno.ilm10b4.hg19’ R package (version 3.13)^47^.

Individual SNP-CpG DNAm association tests were also performed, using the same covariates and p-value corrections as used in the PRS association analyses.

### Gene ontology analysis

Gene ontology analysis was conducted on the MWAS results from GS using the ‘gometh’ function in R package missMethyl^48^. Default settings were used, which include correction for the number of probes per gene. CpGs that showed significant association with depression-PRS at pT<5×10^−8^ in the discovery analysis were selected as CpGs of interest, ‘EPIC’ was chosen for array type and all CpGs included in the analysis were used as the background list. FDR-correction was applied for all analyses.

### Colocalisation analysis

We used Howard et al.’s (2019) MDD GWAS^4^ for MDD-associated SNPs and GoDMC summary statistics for mQTL analysis with PGC studies and GS study removed, which resulted in 32 remaining studies imputed to the 1000 Genome reference panel^13^. We used the package “gwasglue” (version 0.0.0.9000, https://mrcieu.github.io/gwasglue/) to extract SNPs that were +/-1Mb of each of the 102 genome-wide significant, lead SNPs identified in Howard et al. (2019) and then extracted the same SNPs within those regions from the GoDMC mQTL analysis. We used the coloc.abf function with default parameters in the “coloc” package in R (version 5.1.0)^49^ to perform colocalisation analysis for each SNP association. The method tests for five mutually exclusive scenarios in a genetic region: H_0_: there exist no causal variants for either trait; H_1_: there exists a causal variant for trait one only; H_2_: there exists a causal variant for trait two only; H_3_: there exist two distinct causal variants, one for each trait; and H_4_: there exists a single causal variant common to both traits.

### Mendelian randomisation (MR)

Three MR methods, inverse-weighted median (IVW), weighted median (WM) and MR Egger, were used to test for causal effects between DNA methylation and depression using the ‘TwoSampleMR’ R package (version 0.5.6)^50,51^.

GWAS summary statistics for DNAm were from GoDMC and GS. mQTL summary statistics from GoDMC included 32 cohorts with 25,561 participants from European ancestry^13^. The summary statistics were computed using a two-phased design. First, every study performs a full analysis of all candidate mQTL associations, returning only associations at a threshold of p<1×10^−5^. All candidate mQTL associations at p<1×10^−5^ are combined to create a unique ‘candidate list’ of mQTL associations. The candidate list (n=120,212,413) is then sent back to all cohorts, and the association estimates are obtained for every mQTL association on the candidate list. Candidate mQTL associations were meta-analysed using fixed-effect inverse-variance method. Details of the database can be found elsewhere^13^. mQTL summary statistics from GS (N=∼10,000) included a full set of all SNPs with no p-value thresholding. Summary mQTL statistics from GS were generated using the OmicS-data-based Complex Trait Analysis package (https://cnsgenomics.com/software/osca/#eQTL/mQTLAnalysis)^52^. Covariates were consistent with the MWAS for depression-PRS discovery analysis. Further details of the mQTL analysis can be found in the Supplementary Information.

Summary statistics for depression GWAS by Howard *et al*.^33^ were used. A total of 807,553 unrelated, European participants were included in the analysis. Details for the study can be found elsewhere^33^.

GoDMC, GS and depression GWAS samples were mutually exclusive. Individual cohorts that overlapped with the Howard *et al*. depression GWAS and GS were removed from the GoDMC mQTL meta-analysis. Depression GWAS summary statistics from Howard et al (2019) were calculated excluding GS participants^33^. See Supplementary Information for details.

First, we used mQTL summary statistics from GoDMC to identify causal effects from DNA methylation to depression. Second, we used full mQTL summary statistics from GS to replicate the findings and to test the causal effect to DNA methylation and depression bi-directionally. Finally, in contrast to the univariable MR analyses (that is, each risk-outcome pair tested separately), a multi-variable MR analysis was conducted to test for direct causal associations from DNAm at multiple CpGs to MDD, using the mQTL data from GS. Using this method, all CpG probes where there was evidence of a potential causal effect on depression were entered into the two-sample MR analysis simultaneously, in order to prioritise SNPs that showed the strongest independent casual associations with MDD.

Exposures were selected from CpG probes found significantly associated with depression-PRS generated using the p-threshold = 5×10^−8^. As the *MHC* region contains a large number of highly correlated probes, we selected the four independent probes after pruning (r<0.1 for at least two nearest probes, pruning window = 3M base pairs). The probes were further removed from analyses if they did not present in the GoDMC/GS mQTL data or had <5 independent genetic instruments overlapping with the outcome summary statistics. In result, 14 probes were selected for final analyses.

## Results

### Discovery MWAS of depression-PRS in GS

#### Methylome-wide association (MWAS) with depression-PRS at all p thresholds

There were 599 CpG probes significantly associated with depression-PRS with p-threshold (pT) at 5×10^−8^ (p<6.5×10^−8^ to reach significance after Bonferroni correction). In contrast to many other studies that use polygenic risk profiling at different thresholds to predict depression^53^, both the number of significant associations and the effect sizes decreased as PRSs were calculated at increasingly lenient thresholds (Figure 1). For pT of 1×10^−6^, 1×10^−4^ and 0.001, 414, 57 and 17 CpGs were associated with depression-PRS, respectively (Supplementary Figures 1). No significant associations were found for PRS using p-value thresholds greater than or equal to 0.01. Quantile-quantile plot and statistics for genomic inflation factors (ranged from 0.954 to 1.011) can be found in Supplementary Figure 2 and Supplementary Table 4. Results using the depression-PRS calculated using only genome-wide significant variants are presented below.

**Figure 1.**
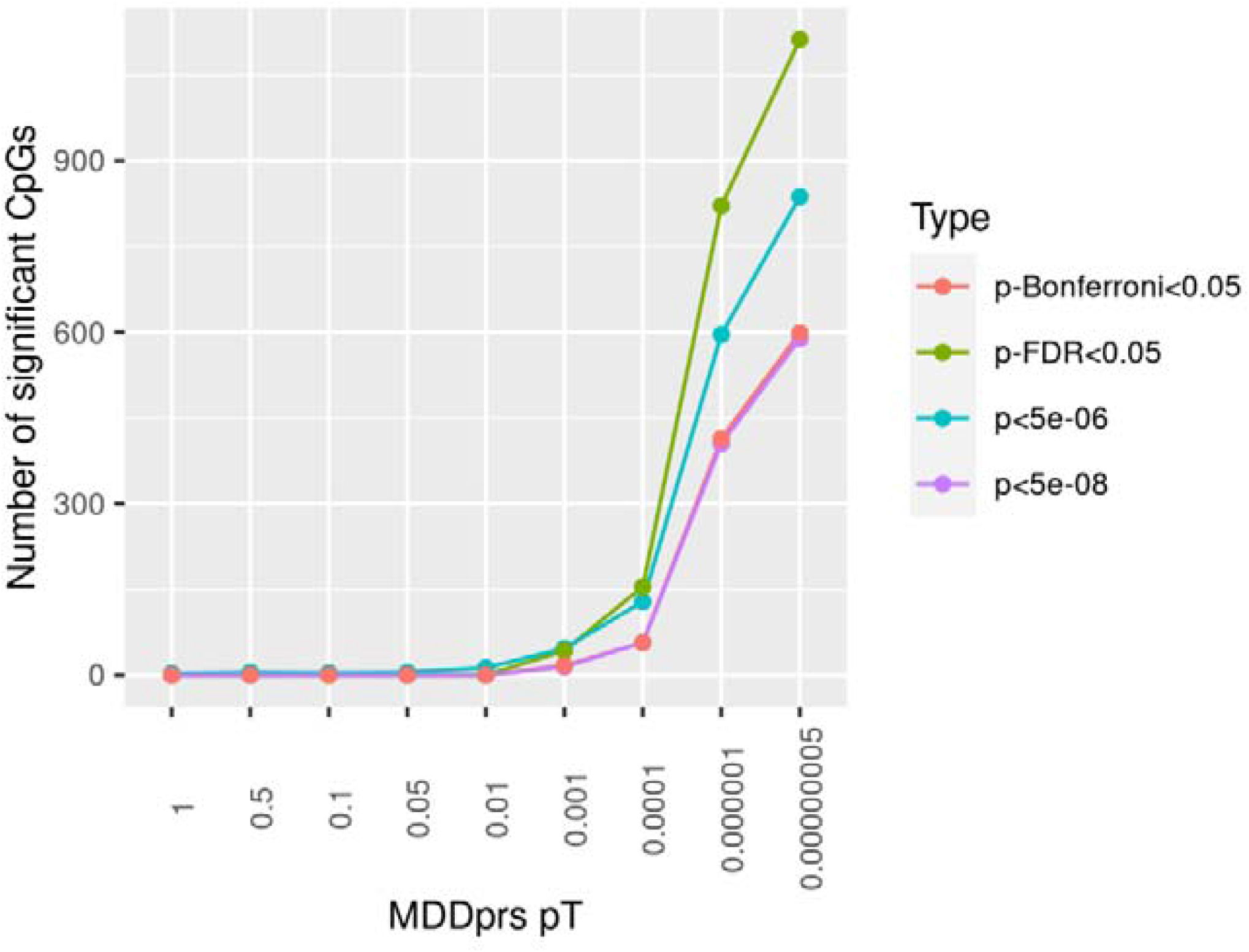
Number of CpG probes associated with polygenic risk scores (PRS) at nine different p-thresholds (pTs) for discovery analysis. X-axis represents the pTs used for generating PRS. Y-axis shows the number of probes significant associated with the given PRS. The four different lines represent four types of methods to define significance.

#### DNAm association with depression-PRS at a GWAS p-value association threshold of 5×10^−8^

The most significant associations of DNAm with depression-PRS were found in the major histocompatibility complex (*MHC*) region (25-35 Mb on Chromosome 6, Figure 2), with 575/599 (96.0%) of significant associations within this region (p_Bonferroni_ ranged from 0.05 to 1.37×10^−111^). The top ten probes that showed the greatest associations are listed in Supplementary Table 5 (all p_Bonferroni_ <2.78×10^−83^). The majority of probes were in Open Sea (47.9%) and CpG shores (28.2%), and they were particularly enriched in the CpG shores compared to non-significant CpGs (18% for non-significant CpGs, χ^2^=41.0, df=1, p=1.53×10^−10^, see Supplementary Table 6 and Supplementary Information). After pruning (r<0.1 for at least two nearest probes, window = 3Mb), four independent CpG probes were identified within the *MHC* region: cg03270340, cg14345882, cg10046620, cg08116408 (all p_Bonferroni_ <2.40×10^−58^). UCSC gene database annotation shows genes that are nearest to the probes are *TRIM27, HIST1H2AI, TRIM40* and *BTN3A2*. See Table 1-2, Supplementary Table 5 and Supplementary Data 1.

**Figure 2.**
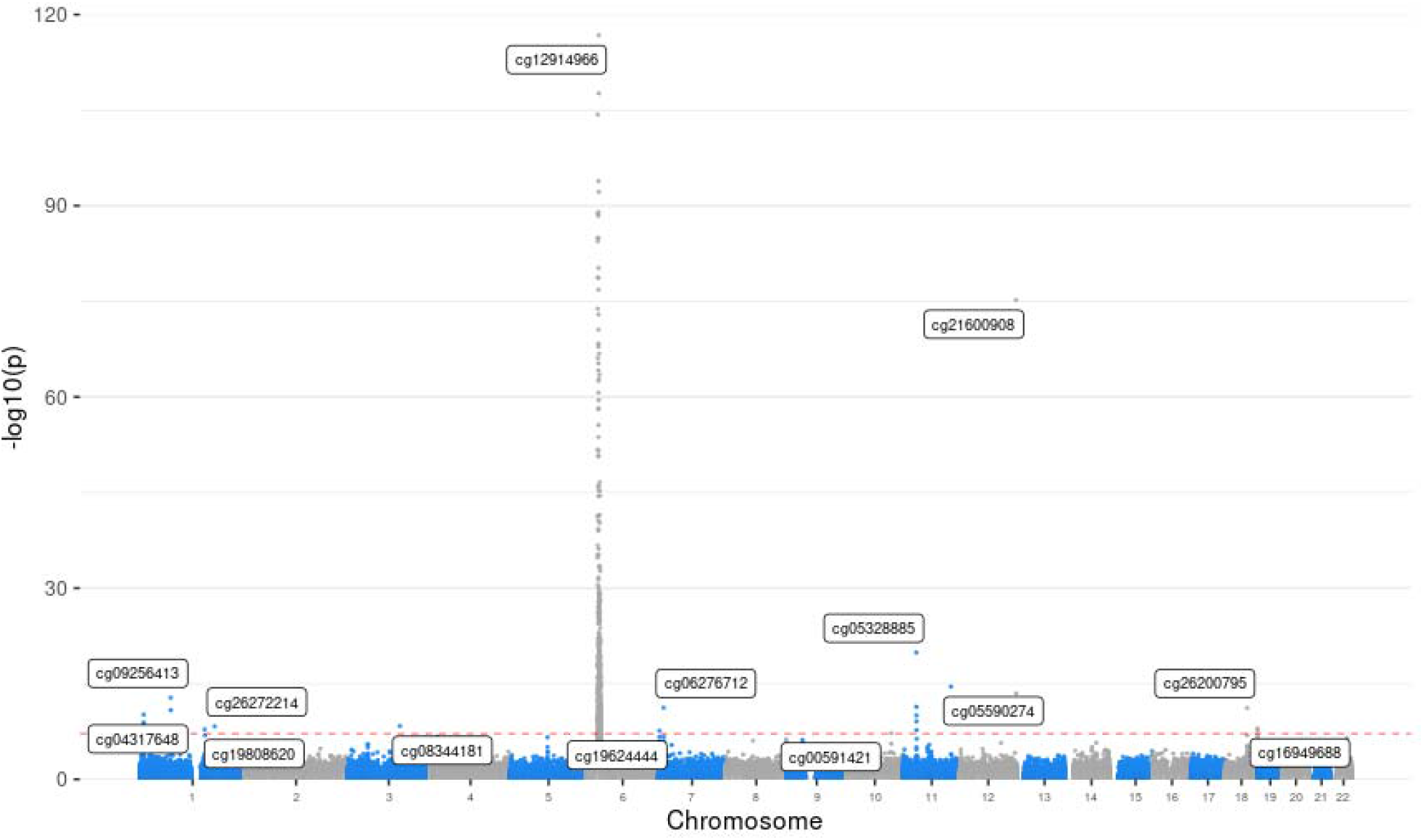
Manhattan plot for the discovery methylome-wide association study (MWAS) for PRS of pT at 5×10^−8^ in Generation Scotland (GS). Each dot represents a CpG probe. X-axis represents the relative position of the probes in the genome. Y-axis represents -log10-transformed p values. The red dashed line represents the significance threshold for Bonferroni correction.

**Table 1.** Results for gene ontology (GO) analysis for the MWAS on PRS at pT 5×10^−8^. Analyses were conducted separately for including and excluding the MHC region. Top ten GO terms are listed in the table. BP=biological process, CC=cellular component and MF=molecular function.

|  | GO ID | Ontology | Term | N | DE | P<sub>DE</sub> | P<sub>FDR</sub> |
| --- | --- | --- | --- | --- | --- | --- | --- |
| With MHC region | GO:0002428 | BP | antigen processing and presentation of peptide antigen via MHC class Ib | 9 | 7 | <1E-322 | <1E-322 |
|  | GO:0002476 | BP | antigen processing and presentation of endogenous peptide antigen via MHC class Ib | 8 | 7 | <1E-322 | <1E-322 |
|  | GO:0002483 | BP | antigen processing and presentation of endogenous peptide antigen | 15 | 8 | <1E-322 | <1E-322 |
|  | GO:0019883 | BP | antigen processing and presentation of endogenous antigen | 22 | 8 | <1E-322 | <1E-322 |
|  | GO:0019885 | BP | antigen processing and presentation of endogenous peptide antigen via MHC class I | 15 | 8 | <1E-322 | <1E-322 |
|  | GO:0042613 | CC | MHC class II protein complex | 14 | 8 | <1E-322 | <1E-322 |
|  | GO:0042611 | CC | MHC protein complex | 23 | 14 | 1.66E-24 | 5.38E-21 |
|  | GO:0042605 | MF | peptide antigen binding | 23 | 11 | 1.97E-17 | 5.61E-14 |
|  | GO:0071556 | CC | integral component of lumenal side of endoplasmic reticulum membrane | 26 | 11 | 3.92E-17 | 8.91E-14 |
|  | GO:0098553 | CC | lumenal side of endoplasmic reticulum membrane | 26 | 11 | 3.92E-17 | 8.91E-14 |
| Without MHC region | GO:0072686 | CC | mitotic spindle | 101 | 2 | 0.002 | 1 |
|  | GO:1902412 | BP | regulation of mitotic cytokinesis | 6 | 1 | 0.002 | 1 |
|  | GO:0021691 | BP | cerebellar Purkinje cell layer maturation | 2 | 1 | 0.002 | 1 |
|  | GO:0021590 | BP | cerebellum maturation | 3 | 1 | 0.002 | 1 |
|  | GO:0021699 | BP | cerebellar cortex maturation | 3 | 1 | 0.002 | 1 |
|  | GO:0045666 | BP | positive regulation of neuron differentiation | 353 | 3 | 0.003 | 1 |
|  | GO:0021578 | BP | hindbrain maturation | 6 | 1 | 0.004 | 1 |
|  | GO:0021626 | BP | central nervous system maturation | 7 | 1 | 0.004 | 1 |
|  | GO:0001093 | MF | TFIIB-class transcription factor binding | 3 | 1 | 0.004 | 1 |
|  | GO:0070369 | CC | beta-catenin-TCF7L2 complex | 3 | 1 | 0.004 | 1 |

**Table 2.** Results for pathway analysis for the MWAS on PRS at pT 5×10^−8^. Analyses were conducted separately for including and excluding the MHC region. Top ten KEGG pathways are listed in the table.

|  | KEGG pathway ID | Description | N | DE | P | P<sub>FDR</sub> |
| --- | --- | --- | --- | --- | --- | --- |
| With MHC region | path:hsa04940 | Type I diabetes mellitus | 41 | 13 | 6.03E-19 | 2.01E-16 |
|  | path:hsa05330 | Allograft rejection | 34 | 12 | 1.17E-18 | 2.01E-16 |
|  | path:hsa05332 | Graft-versus-host disease | 37 | 12 | 2.39E-18 | 2.74E-16 |
|  | path:hsa04612 | Antigen processing and presentation | 66 | 14 | 3.82E-18 | 3.29E-16 |
|  | path:hsa05320 | Autoimmune thyroid disease | 46 | 12 | 2.61E-17 | 1.79E-15 |
|  | path:hsa05416 | Viral myocarditis | 55 | 12 | 8.53E-15 | 4.38E-13 |
|  | path:hsa04145 | Phagosome | 140 | 15 | 8.91E-15 | 4.38E-13 |
|  | path:hsa05150 | Staphylococcus aureus infection | 82 | 10 | 6.89E-12 | 2.96E-10 |
|  | path:hsa05322 | Systemic lupus erythematosus | 112 | 11.5 | 8.87E-12 | 3.39E-10 |
|  | path:hsa04514 | Cell adhesion molecules | 136 | 13 | 2.21E-11 | 7.59E-10 |
| Without MHC region | path:hsa04110 | Cell cycle | 121 | 1 | 0.067 | 1 |
|  | path:hsa04114 | Oocyte meiosis | 114 | 1 | 0.070 | 1 |
|  | path:hsa04514 | Cell adhesion molecules | 118 | 1 | 0.083 | 1 |
|  | path:hsa04914 | Progesterone-mediated oocyte maturation | 86 | 1 | 0.063 | 1 |
|  | path:hsa05166 | Human T-cell leukemia virus 1 infection | 190 | 1 | 0.130 | 1 |
|  | path:hsa05203 | Viral carcinogenesis | 152 | 1 | 0.106 | 1 |
|  | path:hsa00010 | Glycolysis / Gluconeogenesis | 64 | 0 | 1 | 1 |
|  | path:hsa00020 | Citrate cycle (TCA cycle) | 28 | 0 | 1 | 1 |
|  | path:hsa00030 | Pentose phosphate pathway | 25 | 0 | 1 | 1 |
|  | path:hsa00040 | Pentose and glucuronate interconversions | 32 | 0 | 1 | 1 |

Supplementary MWAS were conducted on two additional depression-PRSs to investigate the associations found within the *MHC* region. Two additional PRSs were calculated using (1) the independent genetic risk variants reported in the depression GWAS by Howard *et al*. with a wider pruning window of 3M base pairs and retaining only one variant in the *MHC* region and (2) SNPs located outside of MHC region, respectively. Analysis (1) was conducted to identify if the associations found in the MHC region was due to the additive effect from a large number of genetic variants included in the MHC region. Analysis (2) was conducted to find out if the associations found within the *MHC* region was contributed by genetic variants located *trans* to this region. The number of significant associations found within the MHC region for the PRS calculated using independent genetic risk variants reduced from 599 to 25, at the MDD-GWAS PRS p-value threshold of 5×10^−8^. No CpGs within the *MHC* region were found to be significantly associated with the PRS generated from variants mapping without the MHC region. See Supplementary Figure 3.

Outside of the *MHC* region, 24 probes showed significant associations with depression-PRS estimated across the genome at pT of 5×10^−8^ (p_Bonferroni_ ranged from 0.048 to 4.74×10^−70^). The top ten probes are listed in Supplementary Table 5. Genes mapping near to the top probes were associated with histone deacetylase, DNA binding and transcriptional processes (such as *MAD1L1, TCF4*, RERE and *ZSCAN31*), and neuronal plasticity and growth (for example, *NEGR1*).

The effect sizes for the significant CpG probes showed high correlations between the two data sets (r=0.90), and direction for all significant associations was consistent between sets. For these significant probes, the distance to the nearest depression risk locus was significantly lower than those that were not significant (significant versus not significant: standardised Cohen’s d=0.920, p<1×10^−32^). There were 21.2% of all significant CpGs located outside of 1Mb boundaries of genetic risk loci for depression and outside of the region consisted of SNPs in LD (R^2^>0.1) with the genetic risk loci (see Supplementary Data 1).

### Replication depression-PRS MWAS in LBC1921, LBC1936 and ALSPAC

#### MWAS of depression-PRS on pT of 5×10^−8^ on adult and adolescent samples (LBC1921, LBC1936 and ALSPAC)

We looked at a subset of CpG probes that were significant in the discovery MWAS analysis and found that the standardised effect sizes were correlated between the discovery and replication meta-MWAS of LBC1921, LBC1936 and ALSPAC adults, with (N_probe_=507, r=0.83) or without the probes located in the MHC region (N_probe_=14, r=0.78). There were 98.1% associations found in the discovery MWAS remained in the same direction and 71.8% remained significant after Bonferroni-correction within the replication analysis. See Figure 3.

**Figure 3.**
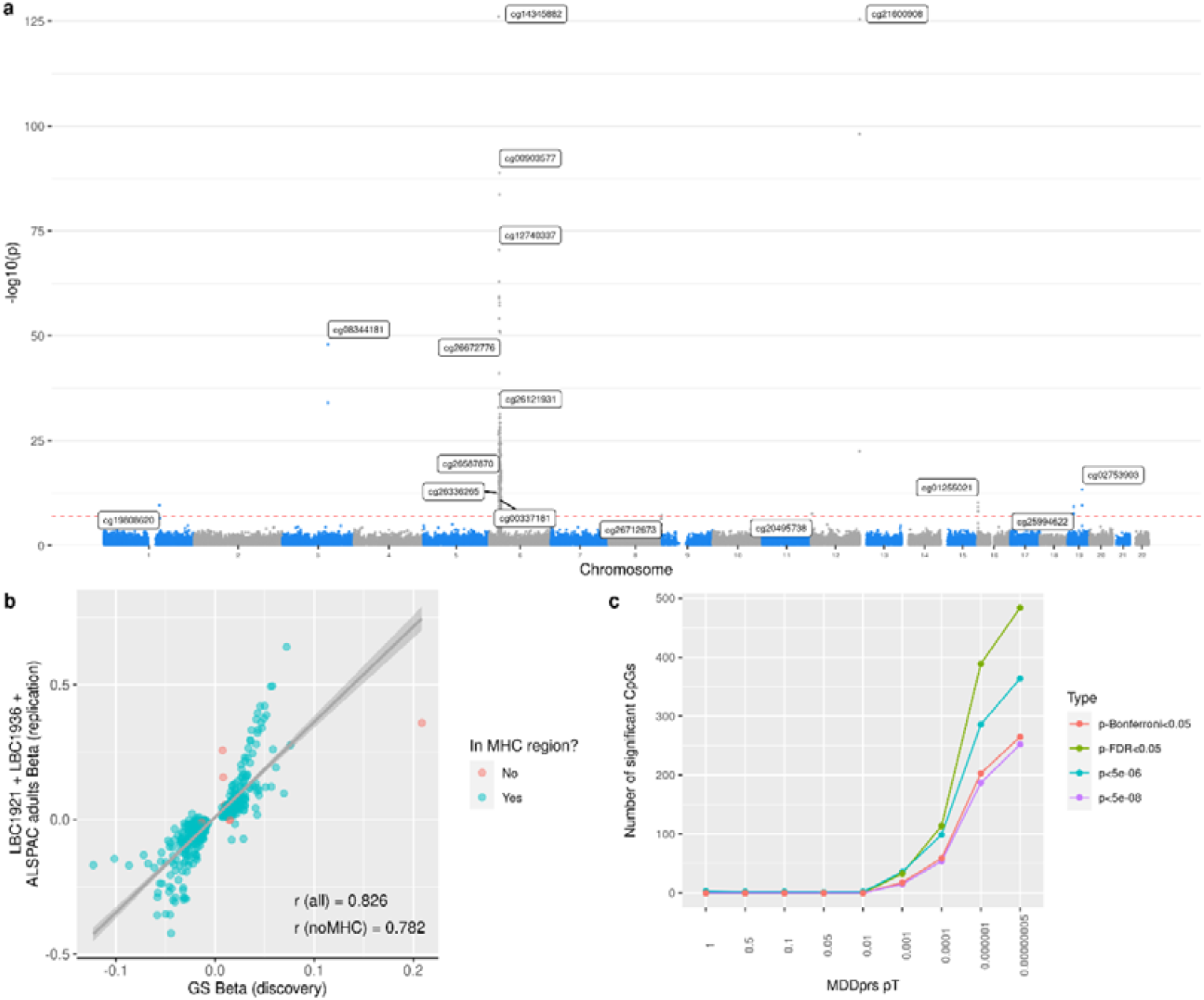
Replication MWAS in Lothian Birth Cohort (LBC) 1921, LBC1936 and Avon Longitudinal Study of Parents and Children (ALSPAC) adults. (a) Manhattan plot for the replication MWAS for PRS of pT at 5×10^−8^. Each dot represents a CpG probe. X-axis represents the relative position of the probes in the genome. Y-axis represents -log10-transformed p values. The red dashed line represents the significance threshold for Bonferroni correction. (b) Number of CpG probes significant associated with polygenic risk scores (PRS) at nine different pTs for replication analysis. X-axis represents the pTs used for generating PRS. Y-axis shows the number of probes significant associated with the given PRS. The four different lines represent four types of methods to define significance. (c) Scatter plot showing the correlation of standardised regression coefficients between the discovery (GS) and replication (LCB1921+LBC1936+ALSPAC adults) analysis. Each dot represents a CpG probe. Probes shown in the figure are those associated with depression-PRS of pT at 5×10^−8^ in the discovery MWAS (in GS). Dots in green represent probes locate in the major histocompatibility complex (*MHC*) regions and those in red represent other probes that locate outside of the MHC region.

Standardised effect sizes for the significant CpG probes found in the discovery MWAS were highly correlated with those in the MWAS on adolescents from ALSPAC (all CpG probes: N_probe_=506, r=0.76; no MHC region: N_probe_=14, r=0.79.

#### MWAS for depression-PRS on all p thresholds on adult samples

Meta-analysis of the methylome-wide association analysis of depression-PRS for replication cohorts (LBC1921, LBC1936 and ALSPAC adults) showed that, for depression-PRS of pT at 5×10^−8^, 1×10^−6^, 1×10^−4^ and 0.001, the number of significant CpG probes were 265, 203, 59 and 18, respectively. Similar to the discovery analysis, no significant associations were found for PRS of pT≥0.01.

A full list of results for replication analysis can be found in Supplementary Data 1 and Supplementary Figures 4-7.

### Pathway enrichment analysis

Gene ontology (GO) terms and KEGG pathways were assessed for the genes associated with depression-PRS of pT at 5×10^−8^. There were 119 enriched GO terms and 29 KEGG pathways significant after FDR correction (all p_FDR_<0.044). The majority of the significant GO terms were associated with antigen processing and the immune response. Enriched KEGG pathways include antigen processing and immune disease, for example, allograft rejection, autoimmune thyroid disease and human T-cell leukemia virus 1 infection. The top ten GO terms and KEGG pathways are listed in Tables 1-2.

A supplementary analysis without probes in the MHC region showed no significant GO terms or KEGG pathways after FDR correction. Top GO terms that were nominally significant include neural development and metabolic process, for example, phosphagen and phosphocreatine metabolic process, cerebellum and cerebellar cortex maturation and cerebellar Purkinje cell layer maturation (p_uncorrected_ ranged from 0.049 to 1.54×10^−3^). No nominally significant KEGG pathway was found (p_uncorrected_>0.063).

### SNP–CpG mapping for the depression risk loci

SNP-CpG probe associations were investigated by conducting MWAS for each of the independent genetic risk loci for depression. There were 6,009 CpG probes that showed significant associations with at least one leading genetic risk variant after Bonferroni correction.

There were 94 of the 96 genetic risk variants tested showed significant *cis* association with CpGs within 1 Mb distance (see Figure 4). There were 88 genetic risk variants (91.6% of all variants tested) that showed *trans* associations outside of their 1Mb window and 82 variants (85.4% of all variants tested) that had *trans* associations with CpGs located on at least one different chromosome.

**Figure 4.**
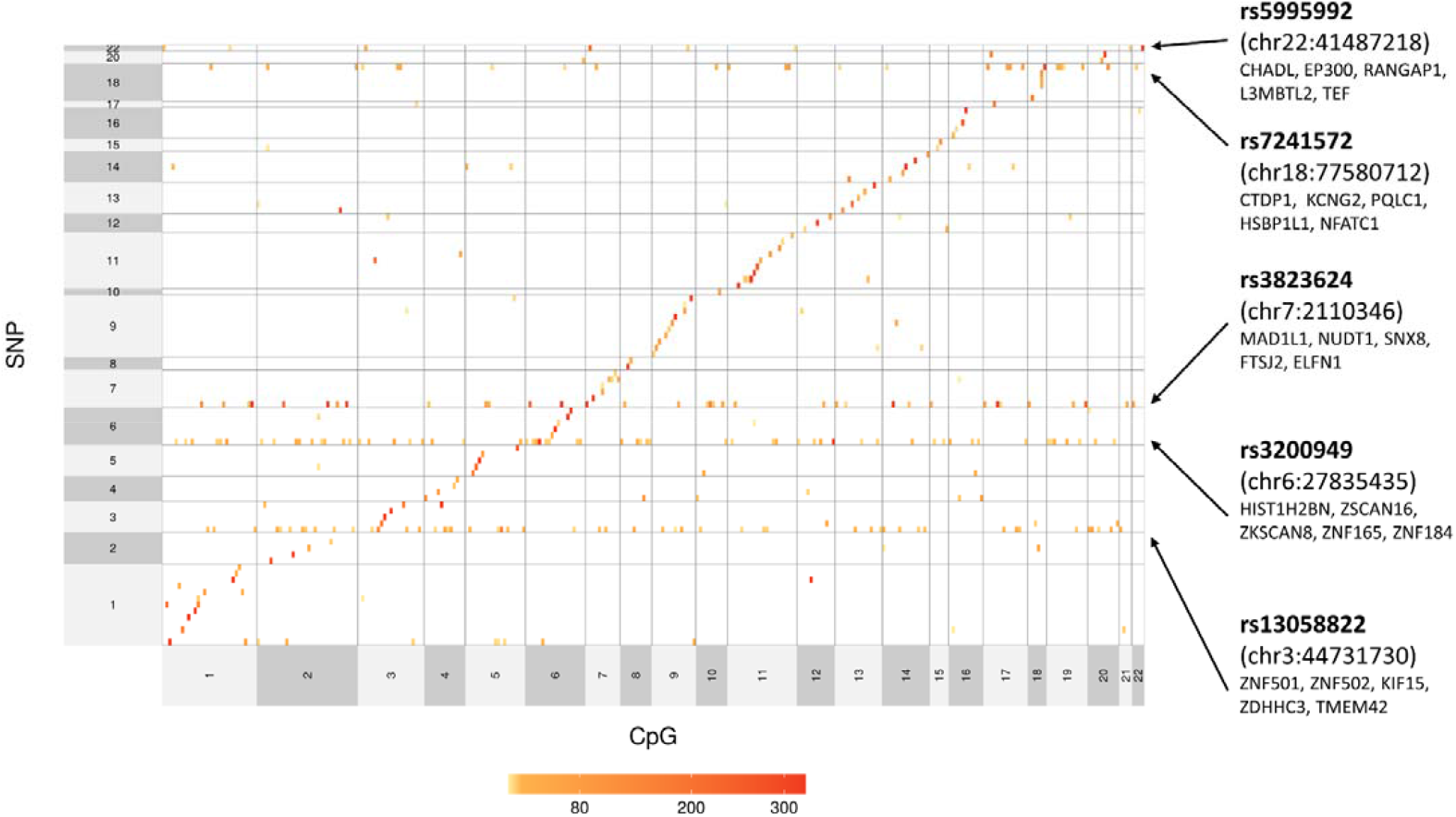
Heatmap showing the SNP-CpG mapping. Each row and column represent a CpG probe and depression risk locus, respectively. Those tests that are not significant after Bonferroni-correction are left blank. For those significant associations, a darker cell represents a higher -log10-transformed p-value. All CpG probes and depression risk loci were categorised based on which chromosome (CHR) they locate. Within each chromosome, probes and SNPs are aligned from left to right or from bottom to top based on their genomic position.

Five genetic loci showed associations with methylation levels at CpGs located in more than half of the distal autosomal chromosomes (see Figure 4). Genes close to these genetic risk variants were involved in, for example, nucleic acid transcription activities, which includes nucleic acid binding (*ZNF179* and *ESR2*), mitotic assembly (*MAD1L1*) and encoding proteins that colocalise with transcription factors (*RERE*). Regional association plots showing genes within 1Mb distance from the five genetic variants can be found in Supplementary Figure 8.

### Colocalisation analysis

We hypothesised that SNPs influencing MDD risk and those influencing DNAm would be shared. Colocalisation analysis, however, indicated that there was no strong evidence (PP_4_>0.8) for shared genetic factors between MDD loci and DNAm. The posterior probability for one region was supportive of a convincing co-localised association signal for both MDD and DNAm in that region (PP_4_=0.710). Within this region, three SNPs were indicated to have a posterior probability of being shared SNPs for the two traits > 0.1: rs73163796=66%, rs73163779=17%, and rs11710605=11%. All three loci colocalised with genetic variation influencing a smoking-associated CpG probe, cg15099418^54^. Supplementary Data 2 contains results for all 102 regions investigated in colocalisation analysis.

### Mendelian randomisation (MR)

#### Discovery MR: causal effect of DNA methylation on depression using GoDMC data

Three probes showed significant causal effect on depression for all three MR methods with consistent direction of effects between methods: cg08344181 on chromosome3, cg17862947 on chromosome 1 and cg17841099 on chromosome 12 (absolute β_IVW_ ranged from 0.042 to 0.103, p_FDR_ ranged from 2.99×10^−47^to 3.60×10^−120^; absolute β_MR-Egger_ ranged from 0.052 to 0.122, p_FDR_ ranged from 0.045 to 1.23×10^−6^, p_FDR_ for MR-Egger intercept ranged from 0.797 to 0.228; absolute β_WM_ ranged from 0.047 to 0.107, p_FDR_ ranged from 1.23×10^−6^ to 2.54×10^−76^, p_FDR_ for Q-statistics ranged from 0.917 to 0.038).

Seven other probes: cg07519229, cg13813247, cg14159747, cg17925084, cg19624444, cg19800032 and cg23275840, showed significant causal effect using the IVW and WM methods (absolute β_IVW_ ranged from 0.019 to 0.046, p_FDR_ ranged from 4.2×10^−3^ to 1.10×10^−8^; β_WM_ ranged from 0.018 to 0.048, p_FDR_ ranged from 1.70×10^−5^ to 4.40×10^−11^, p_FDR_ for Q-statistics ranged from 0.917 to 3.7×10^−28^). Effect sizes were consistent for the above probes were consistent between the IVW and WM methods. No significant causal effect on depression was found using the MR-Egger method for these probes (absolute β_MR-Egger_ ranged from 4.7×10^−7^ to 0.003, p_FDR_ ranged from 0.739 to 0.127). However, the direction of effects remained the same with the IVW and weighted-median methods and the MR-Egger intercept tests showed no evidence of horizontal pleiotropy p_FDR_ ranged from 0.797 to 0.163).

Results are also shown in Figure 5 and Supplementary Figure 9 and Supplementary Table 7.

**Figure 5.**
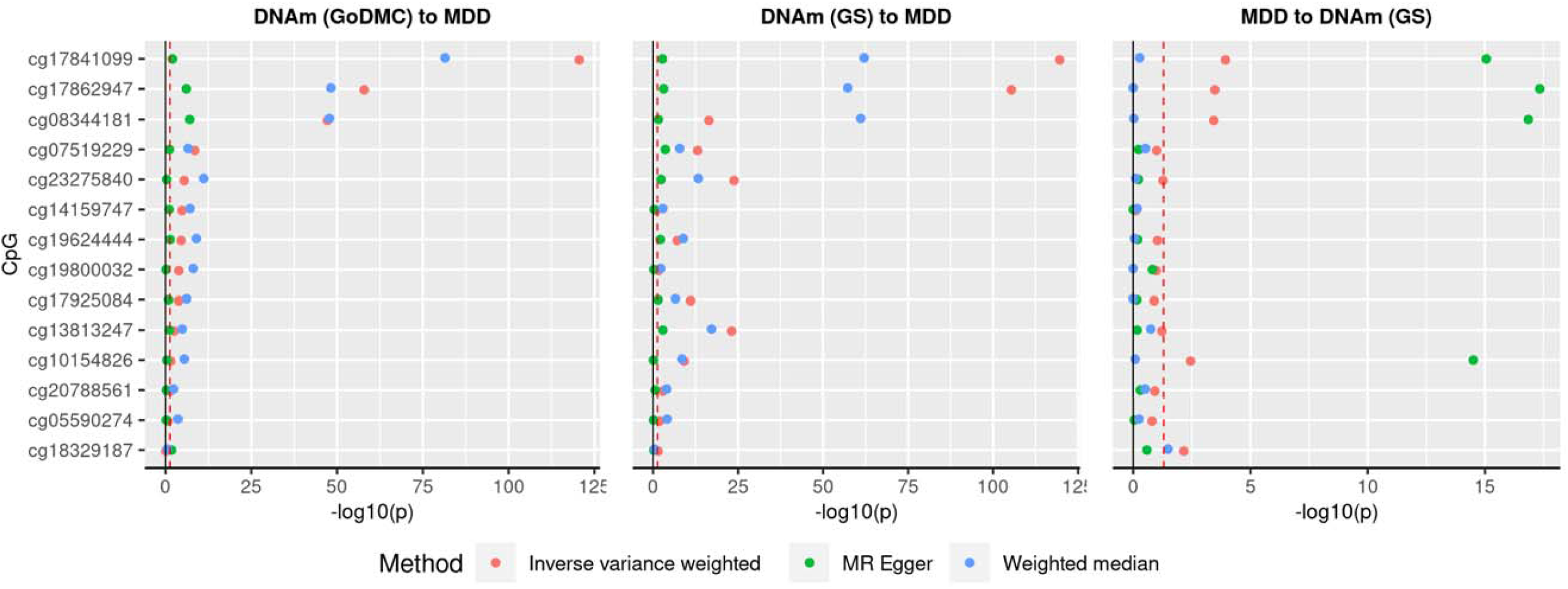
Mendelian randomisation (MR) analysis on DNA methylation and depression using data from the Genetics of DNA Methylation Consortium (GoDMC). (a) Discovery MR testing causal effect of DNA methylation to depression using GoDMC data. (b) Replication MR testing effect of DNA methylation using GS data to depression. (c) MR of reversed directionality testing the causal effect from depression to DNA methylation. X-axes represent p-values for MR analyses. Y-axes represent the individual tests conducted.

#### Replication MR: causal effect of DNA methylation on depression using GS data

All of the causal effects of DNA methylation to depression found in the discovery MR analysis showed consistent direction in the replication analysis and across all three MR methods. For all three MR methods, the effect sizes were highly correlated between discovery and replication analyses (r ranged from 0.855 to 0.934). Eight out of ten significant effects found in the discovery MR analysis were significant for all three MR methods in the replication analyses (absolute β_IVW_ ranged from 0.081 to 1.022, p_IVW-FDR_ ranged from 1.20×10^−7^ to 4.21×10^−119^; absolute β_WM_ ranged from 0.102 to 1.059, p_WM-FDR_ ranged from 1.82×10^−7^ to 2.38×10^−58^). The two other probes showed significant causal effect at two MR methods (cg14159747 significant for MR-Egger and WM and cg19800032 significant for IVW and WM, see statistics in Supplementary Table 3). MR-Egger intercepts were not significantly deviated from 0 for all replication MR (p_FDR_>0.68), and thus showed no evidence of horizontal pleiotropy. See Supplementary Table 8.

#### Multi-variable MR: independent causal effect of DNA methylation on depression using GS data

We next tested for causal associations between DNAm at multiple CpGs from the discovery analysis to MDD. The significant probes were entered into the two-sample MR analysis simultaneously, to identify the set of independent SNPs that showed the strongest and independent casual associations with MDD using the IVW method. Four probes showed causal effects when all CpGs were considered simultaneously. They included cg23275840 on chromosome 4, cg19624444 on chromosome 7, cg17925084 on chromosome 14 and cg17862947 on chromosome 12 (absolute β_IVW_ ranged from 0.077 to 1.023, p_FDR_ ranged from 0.040 to 1.97×10^−6^, see Supplementary Figure 10 and Supplementary Table 9). Genes annotated with these CpG probes are *SLC39A1, CORIN, KLC1* and *MAD1L1*. These genes are involved in signalling receptor binding in the brain and hormonal regulation.

#### MR: causal effects of depression liability on DNA methylation

MR provided evidence of a causal effect of depression liability on DNA methylation for seven CpG probes. Other than the effect to cg09256413 was significant for all three MR methods, effects on other probes were significant for two MR methods (for significant causal effects: absolute β_IVW_ ranged from 0.057 to 0.172, p_IVW-FDR_ ranged from 6.95×10^−3^ to 1.48×10^−5^;absolute β_WM_ ranged from 0.0237 to 0.058, p_WM-FDR_ ranged from 0.046 to 8.24×10^−3^, and absolute β_MR-Egger_ ranged from 0.251 to 6.376, p_MR-Egger-FDR_ ranged from 0.024 to 2.05×10^−23^).

Among all the CpG probes that showed significant causal effects from depression to DNA methylation, only cg17862947 on chromosome 12 showed significant bidirectional effects between depression and DNA methylation (β_IVW_=-0.093, p_FDR_=1.15×10^−4^; β_WM_=-0.008, p_FDR_=0.506; β_MR-Egger_=-0.813, p_FDR_=0.045). However, reversed-MR for this probe showed significant heterogeneity of instrumental effects (p_FDR_ for Q statistics < 1×10^−16^). None of the other CpG probes showed consistently significant bi-directional effect with depression in the discovery and replication MR. See Figure 6, Supplementary Figure 11 and Supplementary Table 10.

## Discussion

Polygenic risk scores (PRS) for genetic risk variants of depression showed wide-spread associations with peripheral blood DNAm across the epigenome in Generation Scotland: Scottish Family Health Study Cohort (GS, N=8,898). The strongest DNAm associations showed highly consistent results in the replication analysis in adults (N=2,049, r_β_ = 0.83) and in adolescents (N=423, r_β_ = 0.76). Significant CpG probes are enriched in immunological processes in the human leukocyte antigen (HLA) system and neuronal maturation and development. Influence from the genetic risk of depression was both local (*cis*) and distal (trans). Five genetic risk loci showed wide-spread *trans* effect across half of the autosomal chromosomes. Finally, using Mendalian randomisation, we found evidence of a mutually causal effect of DNAm on liability to MDD at CpG probes associated with polygenic risk scores for depression.

The probes associated with genetic risk variants for depression map to genes including *TRIM27, BTN3A2* and *HIST1H2AI*. These HLA-related genes have been widely found associated with psychiatric conditions such as schizophrenia and bipolar disorder^6^. Other genes that located outside of the MHC region, such as *BDNF, RERE* and *ZSCAN31*, are associated with neuronal development and guidance of neuronal growth^55^, transcriptional processes^56,57^, as well as other risk factors for depression, for example, obesity, smoking and abnormal physical development^58^.

A large proportion of identified CpG probes were localised in open sea areas and CpG island shores. Previous studies found that probes showed greater individual variations within the CpG island shores^59^, along with higher heritability and greater number of *trans* effect from SNPs^13^. Methylation at these classes of CpGs may be subject to genetic effects to a greater extent than those that map outside such regions. The genes related to the depression genetic risk variants in CpG island shores have previously been linked to DNA, RNA and chromatin binding^60^. Differentially methylation in these regions may reflect a regulatory role for genetic risk variants through influencing DNA methylation in regions adjacent to CpG islands^59^.

Mendelian randomisation provided evidence for a causal effect of methylation levels at CpG probes associated with lead SNPs on depression. After controlling for functional pleiotropy shared between CpG probes, four probes showed an independent causal effect on depression. Genes annotated with these independent probes are associated with phenotypes such as lower total brain volume^61^, excessive C-reactive protein^62^, obesity^63^ and adverse lifestyle factors such as smoking^64^, which are implicated in both patients with depression and those who have been exposed to early environmental risks, such as childhood trauma^65^. These phenotypes have also been shown to have causal effects on depression in previous MR studies^4,5^.

Although evidence was found for bidirectional causal effects between DNA methylation and depression, statistical evidence was stronger for the causal effect from DNA methylation to depression compared to the opposite direction, despite that more genetic variants were used for the reversed causal effect, and thus statistical power was greater (N_SNP_ for DNAm to depression ranged from 5 to 18 and N_SNP_ for depression to DNAm was 112). The highly consistent methylome-wide associations found across adults and adolescents may indicate that early genetic influence on DNAm result in a predominantly directional effect from DNAm to depression^66^.

The present paper utilises large samples with replication analyses yielding highly consistent results. One limitation for interpretating the current findings is that DNA methylation data was collected from blood samples, that may not reflect the most relevant cell types in depression. Nevertheless, studies have shown that the genetic drivers of DNAm have similar effects across multiple cell types^13,67^. The greater accessibility of DNAm from whole blood also has clear sample size and other methodological advantages compared to measures obtained from neural tissue post-mortem, and it is more likely that these measures could be used in future clinical applications. Future studies could further expand the scope by including other cell and tissue types.

In the current study we demonstrate that genome-wide genetic risk variants for depression show wide-spread epigenome-wide DNAm associations both individually and when combined in a risk score. These changes implicate antigen processing and immune system responses and may provide clues to the underlying mechanisms of depression.

## Data Availability

Data from Generation Scotland, Lothian Birth Cohorts and ALSPAC can be obtained with approval. For ALSPAC, the study website contains details of all the data that is available through a fully searchable data dictionary and variable search tool (http://www.bristol.ac.uk/alspac/researchers/our-data/).

## Acknowledgments

Generation Scotland received is currently supported by the Wellcome Trust Investigator Award in Science 01/06/2021 to 31/5/26 ‘Exploiting genomic approaches to identify the environmental basis of depression’. (Reference: 220857/Z/20/Z) to McIntosh AM (PI). The DNA methylation profiling and data preparation was supported by Wellcome Investigator Award 220857/Z/20/Z and Grant 104036/Z/14/Z (PI for both grants is Prof McIntosh AM) and through funding from NARSAD (Ref 27404, Dr Howard DM) and the Royal College of Physicians of Edinburgh (SIM Fellowship, Dr Whalley HC). Genotyping of the GS:SFHS samples was funded by the MRC and Wellcome Trust [104036/Z/14/Z]. Generation Scotland also receives support from the Chief Scientist Office of the Scottish Government Health Directorates [CZD/16/6] and the Scottish Funding Council [HR03006].

We thank the LBC1936 participants and team members who contributed to the LBC1921 and LBC1936 studies. Further study information can be found at https://www.ed.ac.uk/lothian-birth-cohorts. The LBC1921 was supported by the United Kingdom’s Biotechnology and Biological Sciences Research Council (BBSRC), The Royal Society and The Chief Scientist Office of the Scottish Government. The LBC1936 is supported by Age UK (Disconnected Mind project, which supports Dr Harris SE), the Medical Research Council (G0701120, G1001245, MR/M013111/1, MR/R024065/1), and the University of Edinburgh. Methylation typing of LBC1921 and LBC1936 was supported by Centre for Cognitive Ageing and Cognitive Epidemiology (Pilot Fund award), Age UK, The Wellcome Trust Institutional Strategic Support Fund, The University of Edinburgh, and The University of Queensland. Dr Cox SR and Prof Deary IJ were supported by a National Institutes of Health (NIH) research grant R01AG054628, and Dr Cox SR is supported by a Sir Henry Dale Fellowship jointly funded by the Wellcome Trust and the Royal Society (Grant Number 221890/Z/20/Z).

The UK Medical Research Council and Wellcome (Grant Ref: 217065/Z/19/Z) and the University of Bristol provide core support for ALSPAC. A comprehensive list of grants funding is available on the ALSPAC website (http://www.bristol.ac.uk/alspac/external/documents/grant-acknowledgements.pdf). GWAS data was generated by Sample Logistics and Genotyping Facilities at Wellcome Sanger Institute and LabCorp (Laboratory Corporation of America) using support from 23andMe. We are grateful to all the families who took part in this study, the midwives for their help in recruiting them, and the whole ALSPAC team, which includes interviewers, computer and laboratory technicians, clerical workers, research scientists, volunteers, managers, receptionists and nurses. Part of this data was collected using REDCap, see the REDCap website for details https://projectredcap.org/resources/citations/).

## Notes

### Competing Interest Statement

The authors have declared no competing interest.

### Author Declarations

Generation Scotland: Written informed consent for Generation Scotland was obtained for all participants. The study was approved by the NHS Tayside Research Ethics committee (05/s1401/89). Lothian Birth Cohorts: Ethics permission for LBC1921 was obtained from the Lothian Research Ethics Committee (LREC/1998/4/183). Ethics permission for LBC1936 was obtained from the Multi-Centre Research Ethics Committee for Scotland (MREC/01/0/56) and the Lothian Research Ethics Committee (LREC/2003/2/29). ALSPAC: Ethical approval for ALSPAC was obtained from the ALSPAC Ethics and Law Committee and the Local Research Ethics Committees.

