## Supplementary Information for "DNA methylome-wide association study of genetic risk for depression implicates antigen processing and immune responses"

Methylome-wide association analysis of polygenic risk scores for depression

Shen X. et al.

### Methods

#### Additional information for Avon Longitudinal Study of Parents and Children (ALSPAC)

Pregnant women resident in Avon, UK with expected dates of delivery 1st April 1991 to 31st December 1992 were invited to take part in the study. The initial number of pregnancies enrolled is 14,541 (for these at least one questionnaire has been returned or a “Children in Focus” clinic had been attended by 19/07/99). Of these initial pregnancies, there was a total of 14,676 foetuses, resulting in 14,062 live births and 13,988 children who were alive at 1 year of age. When the oldest children were approximately 7 years of age, an attempt was made to bolster the initial sample with eligible cases who had failed to join the study originally. As a result, when considering variables collected from the age of seven onwards (and potentially abstracted from obstetric notes) there are data available for more than the 14,541 pregnancies mentioned above. The number of new pregnancies not in the initial sample (known as Phase I enrolment) that are currently represented on the built files and reflecting enrolment status at the age of 24 is 913 (456, 262 and 195 recruited during Phases II, III and IV respectively), resulting in an additional 913 children being enrolled. The phases of enrolment are described in more detail in the cohort profile paper and its update (see footnote 4 below). The total sample size for analyses using any data collected after the age of seven is therefore 15,454 pregnancies, resulting in 15,589 foetuses. Of these 14,901 were alive at 1 year of age. A 10% sample of the ALSPAC cohort, known as the Children in Focus (CiF) group, attended clinics at the University of Bristol at various time intervals between 4 to 61 months of age. The CiF group were chosen at random from the last 6 months of ALSPAC births (1432 families attended at least one clinic). Excluded were those mothers who had moved out of the area or were lost to follow-up, and those partaking in another study of infant development in Avon.

Study data were collected and managed using REDCap electronic data capture tools hosted at the University of Bristol^1^. REDCap (Research Electronic Data Capture) is a secure, web-based software platform designed to support data capture for research studies.

#### Covarying confounders in the methylome-wide association study (MWAS)

##### Generation Scotland: **Scottish Family Health Study (GS)**

Technical confounders were pre-corrected. This was achived by residualising m-values against genetic relationship matrix (GRM), batch and estimated cell proportions. GRM was generated using Plink2.0^2^ and was corrected in wave 1 only, as participants were unrelated in wave 2. Other covariates were fitted in the MWAS regression model, which include: age, sex, pack years, ever smoked tobacco and the first 20 principal components of the methylation data.

Ever smoked tobacco was an ordinal variable. Participants were asked to choose from one of the following responses: ‘Yes, currently smoke’ (= 3), ‘Yes, but stopped within the past 12 months’ (= 2), ‘Yes, but stopped for more than 12 months ago’ (= 1) and ‘No, never smoked’ (= 0). This variable was set as factor in all analyses using R.

Principal components of the methylation data was derived using the ‘FactoMineR’ R package (version 2.4)^3^. This step was conducted on the m-values pre-corrected for technical confounders.

##### Lothian Birth Cohort (LBC) 1921 and LBC 1936

All participants were unrelated within each cohort itself and between the two cohorts. All covariates were fitted in the MWAS regression model, which include: estimated cell proportions, batch, age, sex, ever smoked tobacco and the first 20 principal components of the methylation data.

Ever smoked tobacco variable was ordinal and thus was set as factor in all analyses. Responses include: ‘Current smoker’ (= 2), ‘Previous smoker’ (= 1) and ‘Never smoked’ (= 0).

Principal components of the methylation data was derived using the same method as GS.

##### ALSPAC

MWAS was conducted on parents and youth respectively. For each of the MWAS, only unrelated participants were included in the analyses. Procedures for selecting unrelated sample can be found in^4^. Covariates that were fitted in the MWAS analysis include: estimated cell proportions, age, sex, ever smoked tobacco, the first 15 variables derived from surrogate variable analysis and the first 10 genetic principal components.

Ever smoked tobacco was asked around the time of blood draw for DNAm processing. Participants were asked to choose from either smoked (= 1) or not smoked (= 0).

#### Meta-analysis on MWAS results

Meta-analysis was conducted using METAL (version published on 25/03/2011)^5^ using the fixed-effect inverse-variance method.

For each individual analyses in the discovery and replication MWAS, meta-analysis was conducted to obtain an overall summary statistics of MWAS results. In the discovery analysis, MWAS on set 1 and set 2 GS data was conducted separately and then meta-analysed. In the replication analysis, MWAS was conducted on the two LBC cohorts together and ALSPAC adults. Summary statistics for replication analyses were then meta-analysed.

A final meta-analysis on the discovery and replication MWAS (LBC cohorts and ALSPAC adults) was conducted and presented in the Supplementary Information.

As there was extensive control for relatedness and population structure for MWAS in all cohorts, genomic control correction was not included in the analyses.

#### Enrichment for CpG genomic positions

Enrichment test for CpG genomic positions was conducted to compare the numbers of CpG probes landed in each category for genomic position between those significantly associated with MDD PRS at pT=5⨉10^-8^ and those that did not show significant associations.

Chi-square test for enrichment was conducted using the ‘chisq.test’ function in R (‘stats’ package, version 3.6.2). Pearson’s Chi-squared test with Yates’ continuity correction was used for the analysis.

CpG positions were categorised using the ‘Relation_to_Island’ column from the annotation object derived using the ‘IlluminaHumanMethylationEPICanno.ilm10b4.hg19’ R package (version 3.13). There were six categories according to the UCSC classification of CpG islands: Island (within a CpG island), N-shore (in the 2 kb upstream region of CpG island boundaries), S-shore (in the 2 kb downstream region of CpG island boundaries), N-shelf (in the 2-4kb upstream region of CpG island boundaries), S-shelf (in the 2-4kb downstream region of CpG island boundaries) and Open Sea.

For each category, a chi-square test was performed on a 2⨉2 matrix of counts for CpG probes. Rows of the matrix are posistions of the target analysis and other positions, and columns are significant CpGs and non-significant CpGs. For example, when enrichment of the ‘Island’ position was tested, the matrix is consisted of counts (N) for significant CpGs within an Island (row 1, column 1), non-significant CpGs within an Island (row 1, column 2), significant CpGs outside of any Island (row 2, column 1) and non-significant outside of any Island (row 2, column 2).

Results can be found in Supplementary Table 6.

#### Statistics for methylation quantitative trait loci (mQTL)

##### **Genetics of DNA Methylation Consortium** (GoDMC)

GoDMC([<http://www.godmc.org.uk/cohorts.html>](http://www.godmc.org.uk/cohorts.html)) was established with the view of bringing together researchers with an interest in studying the genetic basis of DNA methylation variation, to consolidate as many resources and expertise as possible and thereby expedite this field of research^6^[.](https://www.medrxiv.org/content/10.1101/2020.09.01.20180406v1%7D.) The initial release of their findings consists of mQTL associations based on a sample size of 27,750 individuals. For the present study, LBC 1921, LBC 1936, GSK, and Brisbane Systems Genetics Study were removed from the mQTL meta-analysis as they were also included in the MDD GWAS by Howard *et al.*^7^. Details for the cohorts can be found at the GoDMC website . As a result, a total of ~25,000 participants were left in the meta-analysis, with no overlapping participants with GS and no overlapping cohort with the MDD GWAS^7^.

*Genotype data*

Genotype data of all autosomes and chromosome X (if available) was imputed to 1000G and above using hg19/build37. Genotype data was filtered on an info score of 0.8 and a minor allele frequency (MAF) of 0.01. Genotype data was converted to best-guess data without a probability cut-off.

*DNA methylation data*

DNA methylation was measured in whole blood or cord blood using Illumina 450k or EPIC Beadchips in at least 100 European individuals. Normalized beta values were used, preferable normalized with the R package ‘meffil’ (version 1.1.1)8 Most analysts used meffil to quality control and normalize the DNA methylation data using functional normalization. Protocols can be found here: <https://github.com/perishky/meffil/wiki>.

A github pipeline was implemented to run the analyses locally ([https://github.com/MRCIEU/godmc).](https://github.com/MRCIEU/godmc).) For the genotype data, several standard sample QC steps were performed including a sex check, removal of samples with >5% missingness, and the identification and exclusion of ancestry outliers. In datasets of ostensibly unrelated individuals, those that were found to be related (identity by state > 0.125) were excluded.

The pipeline then residualised the normalized methylation betas by replacing outliers that were 10 standard deviations from the mean (3 iterations) with the probe mean, rank transforming the normalized beta values and regressing out age, sex, predicted cell counts, predicted smoking, genetic principal components and non-genetic methylation principal components. In family-based cohorts, genetic relatedness matrices were constructed and relatedness adjusted for using the ‘GRAMMAR’ approach. Genomic lambdas were checked by performing a GWAS of probe cg07959070. These residualised methylation measurements were used in all analyses.

*Association analysis*

First, every study performed a full analysis of all candidate mQTL associations, returning only associations at a threshold of p<1e-5. All candidate mQTL associations at p<1e-5 were combined to create a unique ‘candidate list’ of mQTL associations. In total, 102,965,711 candidate mQTL associations in *cis* (p<1⨉10^-5^, SNP located within 1Mb of the methylation site) and 710,638,230 candidate mQTL associations in *trans* were identified in at least one dataset. To avoid computational burden, we included *cis* associations found in at least one dataset and *trans* associations in at least two datasets. The candidate list (n=120,212,413) was then sent back to all cohorts and the association estimates obtained for every mQTL association on the candidate list. Meta analyses were run using a modified version of METAL using fixed-effect inverse-variance method^5^.

#### GS

Analysis of mQTL was conducted on DNAm data in GS for set 1 and set 2 separately. The OmicS-data-based Complex trait Analysis package was used for deriving mQTL summary statistics (<https://cnsgenomics.com/software/osca/>)^9^. DNAm data and covariates were kept consistent with the MWAS. Genetic data used for the mQTL analysis was also used for calculating polygenic risk scores for depression. Finally, meta-analysis between set 1 and set 2 was conducted on the mQTL summary statistics for each CpG probe.

###### Supplementary Figure 1 (a-f). Manhattan plots for discovery MWAS on GS. Panels (a-i) are for PRSs at p thresholds 5⨉10_-8_, 1⨉10_-6_, 1⨉10_-4_, 1⨉10_-3_, 0.005, 0.01, 0.05, 0.1, 0.2, 0.5 and 1.

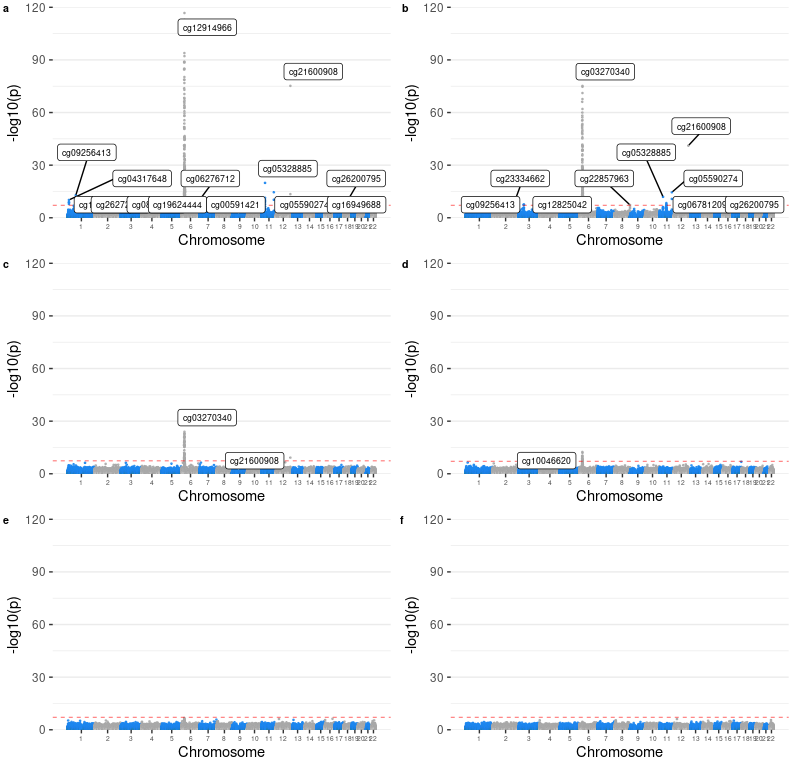

###### Supplementary Figure 1 (g-j). Manhattan plots for discovery MWAS on GS. Panels (a-i) are for PRSs at p thresholds 5⨉10_-8_, 1⨉10_-6_, 1⨉10_-4_, 1⨉10_-3_, 0.005, 0.01, 0.05, 0.1, 0.2, 0.5 and 1.

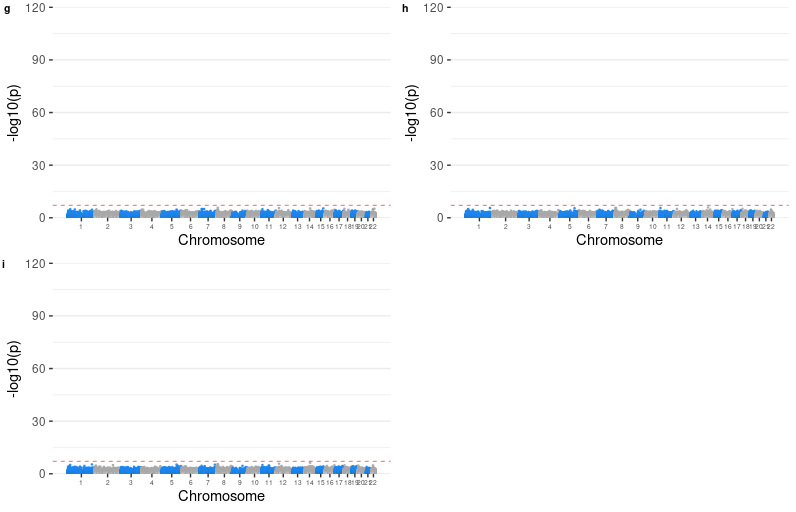

###### Supplementary Figure 2. Quantile-quantile plots for methylome-wide association studies (MWAS) of polygenic risk scores (PRS) for depression at p threshold of 5e-8 on Generation Scotland: Scottish Family Health Study (GS).

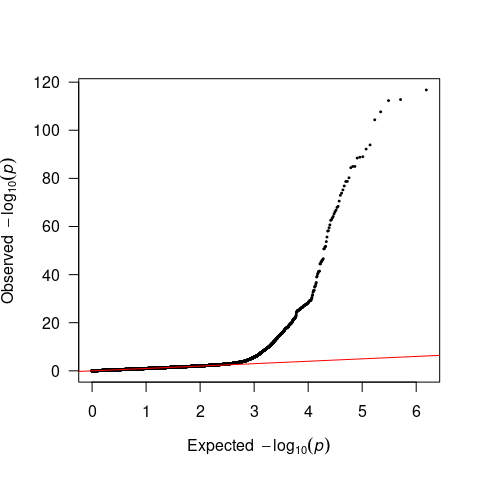

###### Supplementary Figure 3. Supplementary MWAS investigating the MHC region. (a) MWAS for PRS calculated using the leading genetic risk variants. (b) MWAS for PRS calculated using genome-wide significant variants located outside of the MHC region.

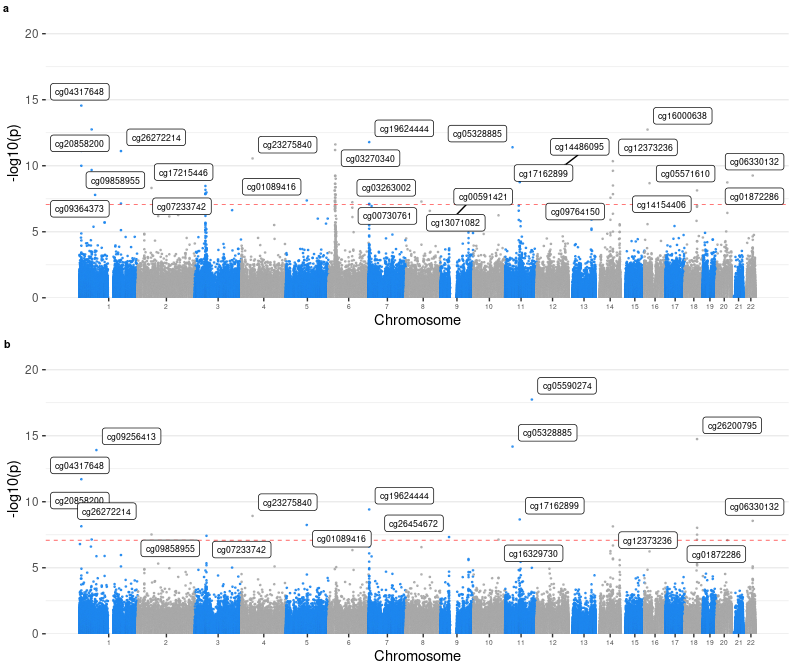

###### Supplementary Figure 4 (a-f). Manhattan plots for replication MWAS on LBC 1921, LBC 1963 and ALSPAC adults. Panels (a-i) are for PRSs at p thresholds 5⨉10_-8_, 1⨉10_-6_, 1⨉10_-4_, 1⨉10_-3_, 0.005, 0.01, 0.05, 0.1, 0.2, 0.5 and 1.

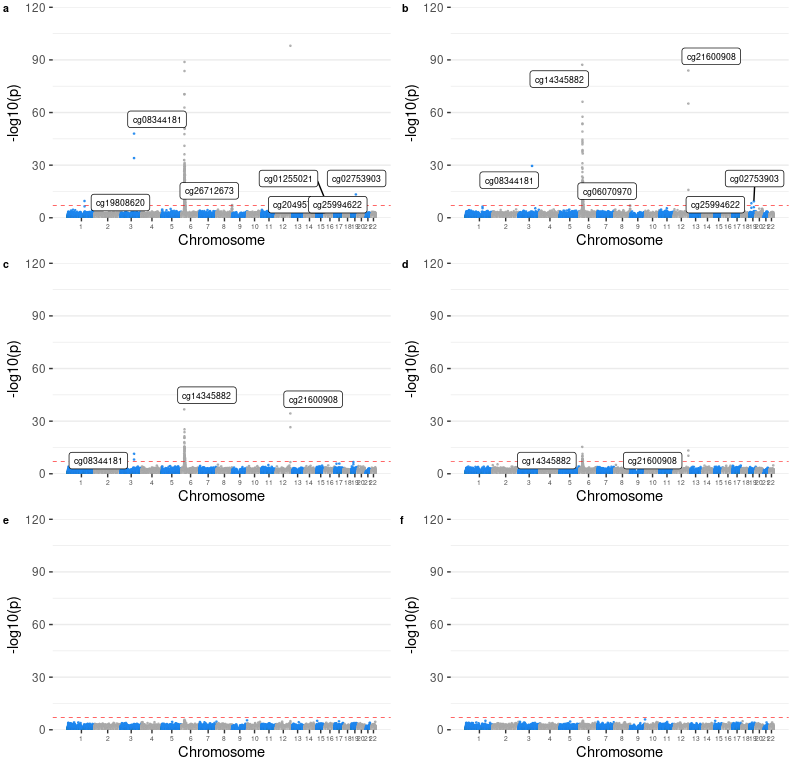

###### Supplementary Figure 4 (g-j). Manhattan plots for replication MWAS on LBC 1921, LBC 1963 and ALSPAC adults. Panels (a-i) are for PRSs at p thresholds 5⨉10_-8_, 1⨉10_-6_, 1⨉10_-4_, 1⨉10_-3_, 0.005, 0.01, 0.05, 0.1, 0.2, 0.5 and 1.

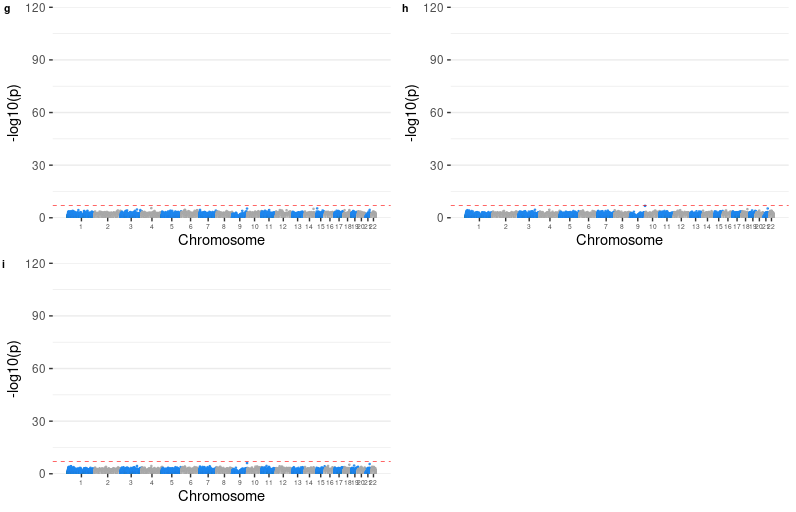

###### Supplementary Figure 5. Quantile-quantile plots for replication MWAS of PRS for depression at p threshold of 5e-8 on Lothian Birth Cohort (LBC) 1921, LBC 1936 and Avon Longitudinal Study of Parents and Children (ALSPAC) adults.

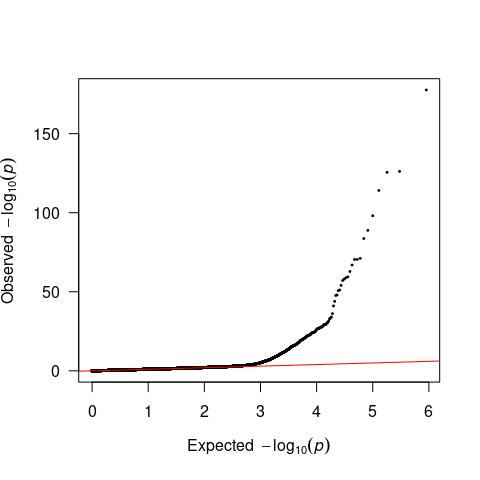

###### Supplementary Figure 6 (a-f). Manhattan plots for replication MWAS ALSPAC children. Panels (a-i) are for PRSs at p thresholds 5⨉10_-8_, 1⨉10_-6_, 1⨉10_-4_, 1⨉10_-3_, 0.005, 0.01, 0.05, 0.1, 0.2, 0.5 and 1.

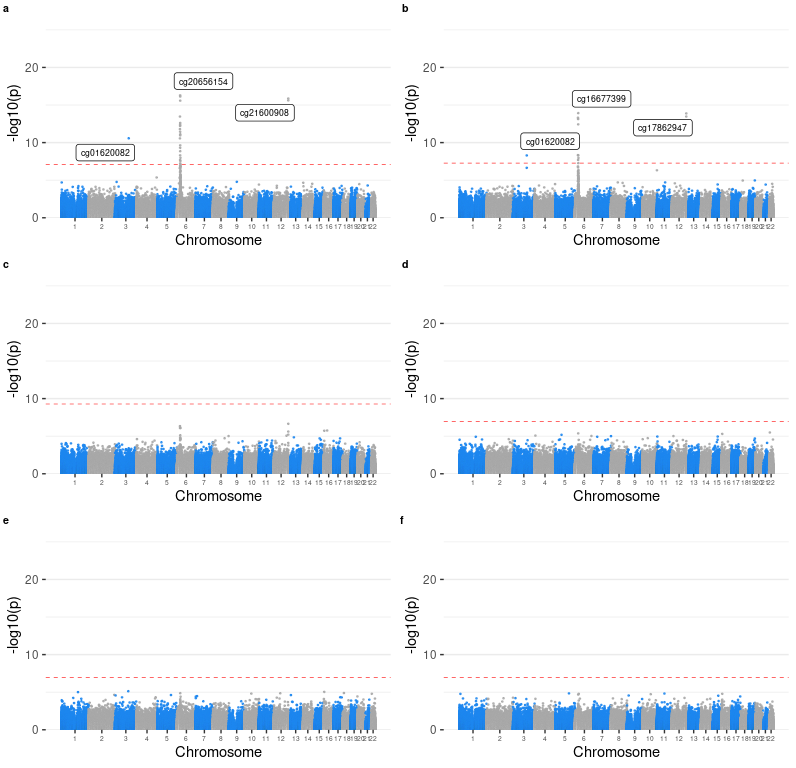

###### Supplementary Figure 6 (g-j). Manhattan plots for replication MWAS ALSPAC children. Panels (a-i) are for PRSs at p thresholds 5⨉10_-8_, 1⨉10_-6_, 1⨉10_-4_, 1⨉10_-3_, 0.005, 0.01, 0.05, 0.1, 0.2, 0.5 and 1.

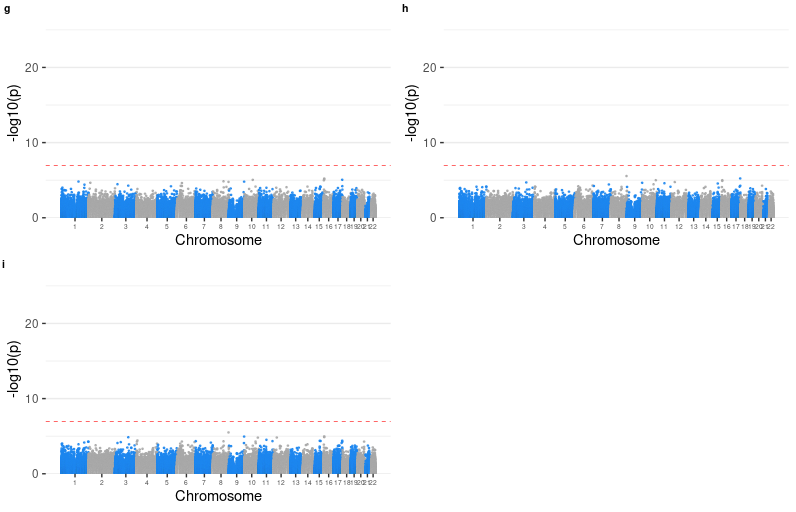

###### Supplementary Figure 7. Quantile-quantile plots for replication MWAS of PRS for depression at p threshold of 5⨉10_-8_ on ALSPAC children.

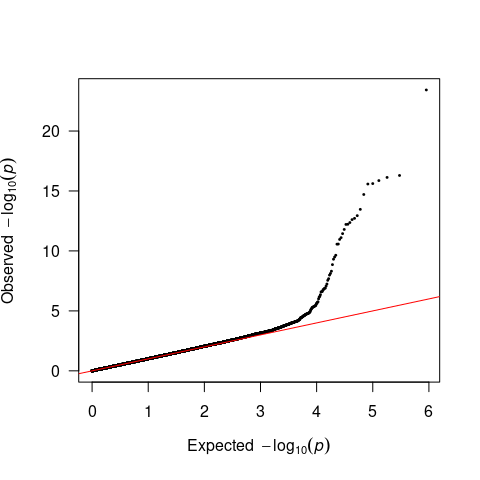

###### Supplementary Figure 8. Regional association plots for the genetic loci showed associations with methylation levels at CpGs located in more than half of the distal autosomal chromosomes (window = 1Mb).

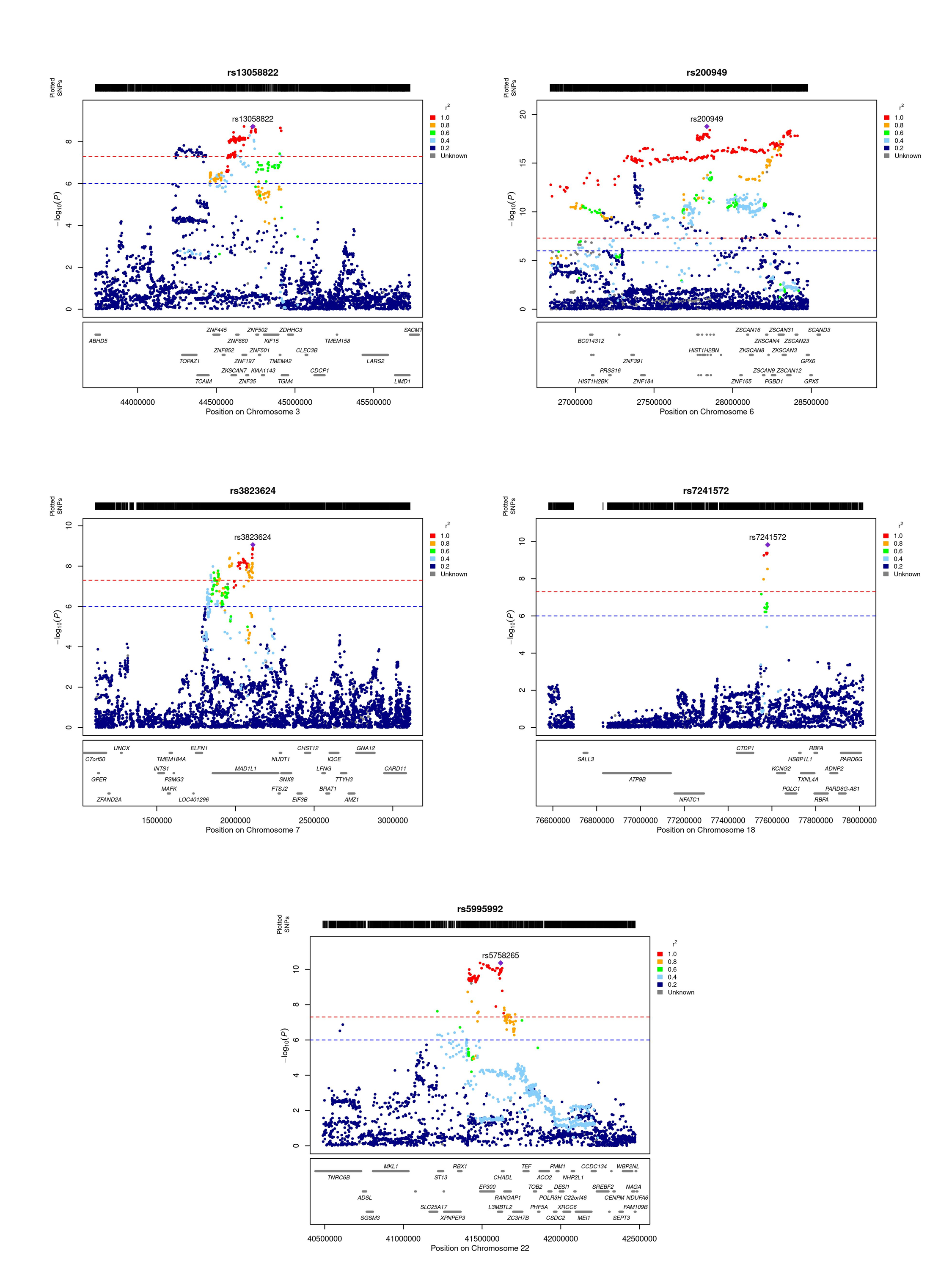

###### Supplementary Figure 9. Scatter plot for discovery Mendelian randomisation (MR) of DNA methylation (DNAm) to depression (a-i).

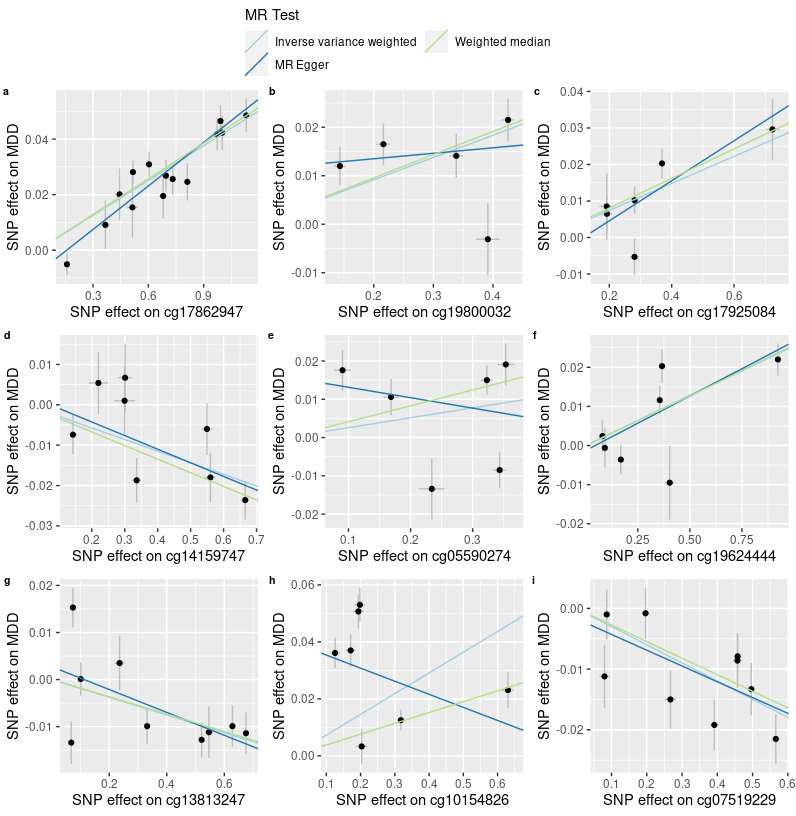

###### Supplementary Figure 9. Scatter plot for discovery Mendelian randomisation (MR) of DNA methylation (DNAm) to depression (j-n).

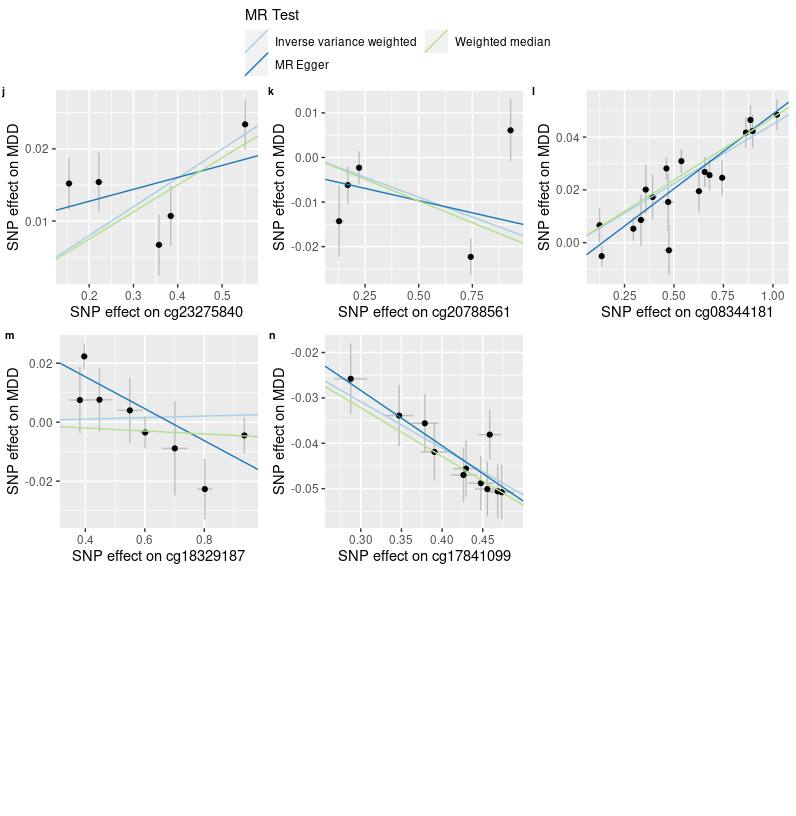

###### Supplementary Figure 10. Scatter plot for replication MR of DNAm to depression (a-i).

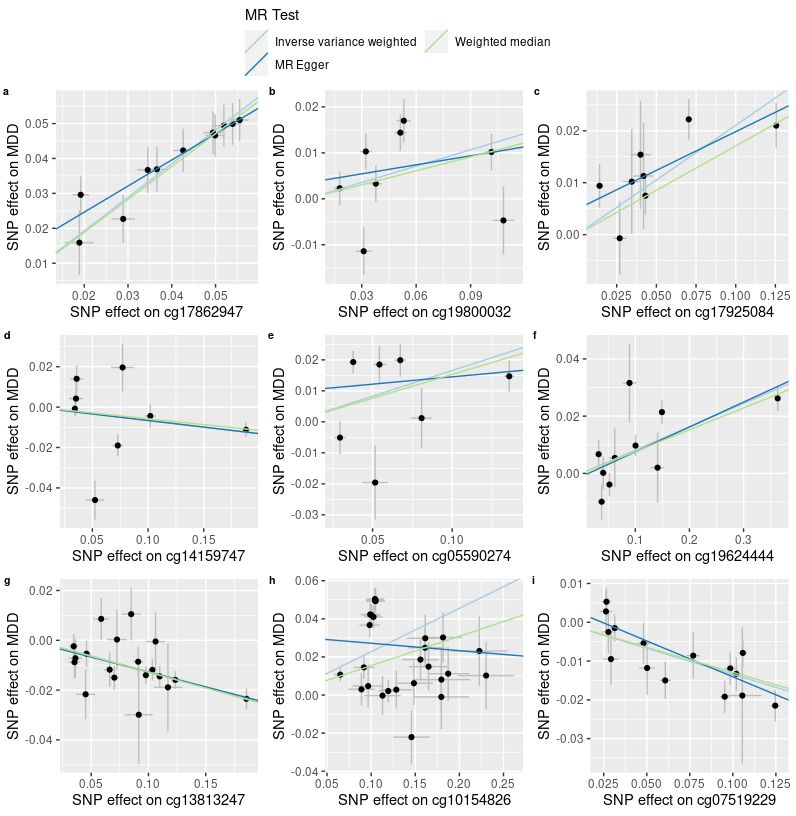

###### Supplementary Figure 10. Scatter plot for replication MR of DNAm to depression (j-n).

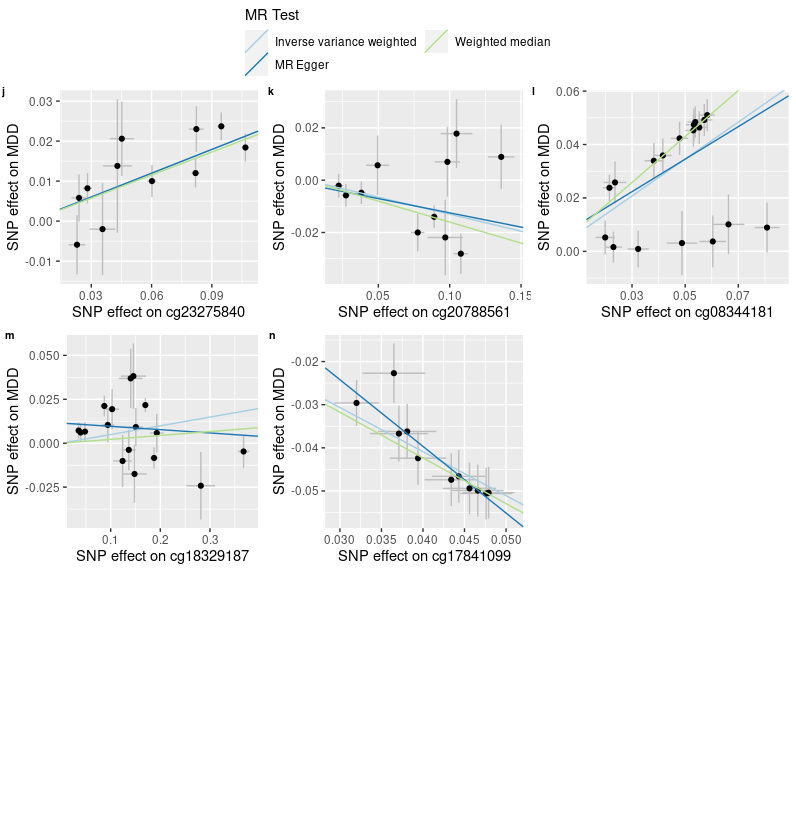

###### Supplementary Figure 11. Scatter plot for MR of liability of depression to DNAm (a-i).

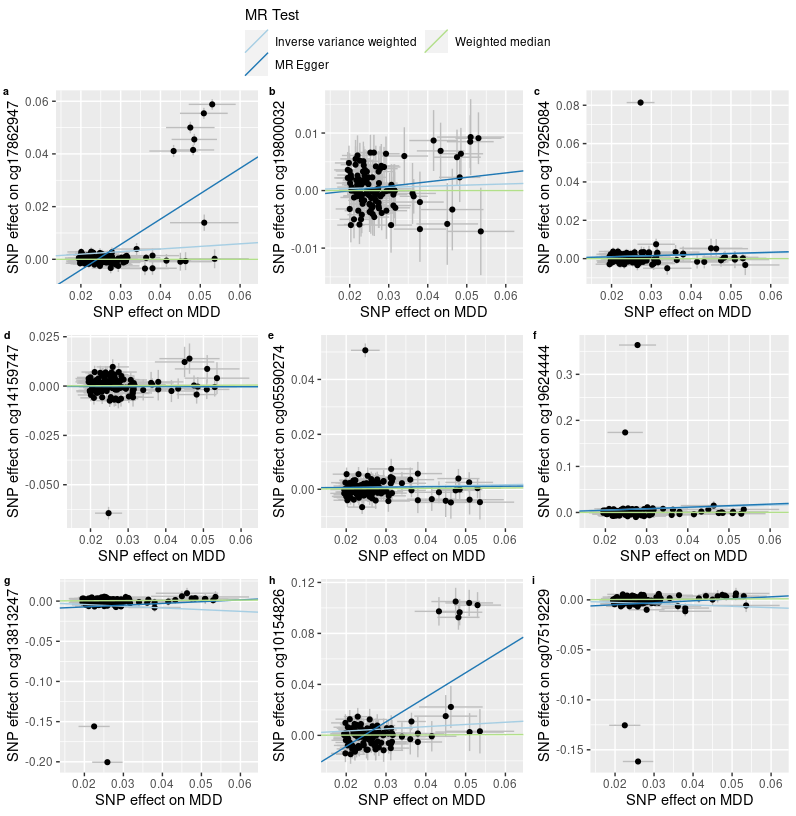

###### Supplementary Figure 11. Scatter plot for MR of liability of depression to DNAm (j-n).

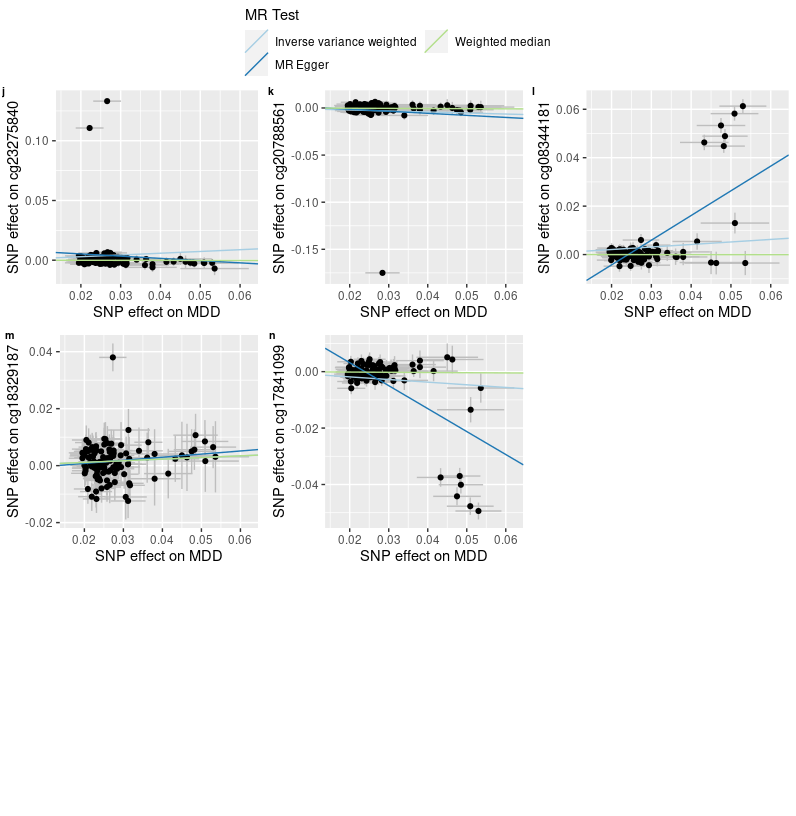

###### Supplementary Table 1. Number of genetic variants used for calculating PRSs. pT = p threshold used for calculating PRS, N_SNP_ = total number of SNPs used for calculating PRS, N_noMHC_ = number of SNPs outside of the MHC region, N_MHC_ = number of SNPs within the MHC region.

| pT | N_SNP_ | N_noMHC_ | N_MHC_ |
| --- | --- | --- | --- |
| 5e-08 | 127 | 118 | 9 |
| 1e-05 | 615 | 602 | 13 |
| 1e-04 | 1457 | 1441 | 16 |
| 1e-03 | 4199 | 4176 | 23 |
| 0.01 | 14578 | 14530 | 48 |
| 0.05 | 38284 | 38185 | 99 |
| 0.1 | 58867 | 58742 | 125 |
| 0.5 | 151226 | 150977 | 249 |
| 1 | 199432 | 199113 | 319 |

###### Supplementary Table 2. Association between PRSs and prevalence MDD. PRS pT = p threshold used for selecting SNPs for calculating PRS.

| PRS pT | Beta | SE | p | R2 (%) |
| --- | --- | --- | --- | --- |
| 5e-08 | 0.114 | 0.033 | 5.52e-04 | 0.269 |
| 1e-05 | 0.157 | 0.032 | 1.28e-06 | 0.496 |
| 1e-04 | 0.208 | 0.032 | 4.80e-11 | 0.926 |
| 1e-03 | 0.236 | 0.032 | 1.40e-13 | 1.149 |
| 0.01 | 0.253 | 0.032 | 2.07e-15 | 1.332 |
| 0.05 | 0.323 | 0.033 | 3.02e-23 | 2.086 |
| 0.1 | 0.326 | 0.032 | 5.38e-24 | 2.161 |
| 0.5 | 0.294 | 0.032 | 8.20e-20 | 1.752 |
| 1 | 0.288 | 0.032 | 3.78e-19 | 1.688 |

###### Supplementary Table 3. Association between PRSs and DNAm-estimated white-blood cell proportions.

|  | CD8T | | CD4T | | NK | | Bcell | | Mono | | Gran | |
| --- | --- | --- | --- | --- | --- | --- | --- | --- | --- | --- | --- | --- |
| PRS pT/MDD | Beta | p | Beta | p | Beta | p | Beta | p | Beta | p | Beta | p |
| 0.00000005 | 0.78 | 0.079 | -1.316 | 0.054 | 0.238 | 0.594 | -0.162 | 0.63 | 0.42 | 0.113 | 0.039 | 0.97 |
| 0.000001 | 0.525 | 0.476 | -1.236 | 0.275 | -0.039 | 0.958 | -0.315 | 0.572 | 0.27 | 0.54 | 0.796 | 0.644 |
| 0.0001 | -1.012 | 0.652 | 0.383 | 0.912 | 2.06 | 0.36 | 0.429 | 0.8 | 0.95 | 0.478 | -2.811 | 0.593 |
| 0.001 | -7.709 | 0.085 | 6.729 | 0.329 | 1.34 | 0.765 | -1.442 | 0.67 | 2.603 | 0.331 | -1.521 | 0.885 |
| 0.01 | -11.225 | 0.264 | 15.929 | 0.303 | 8.285 | 0.411 | -5.734 | 0.451 | -0.025 | 0.997 | -7.229 | 0.759 |
| 0.05 | -10.981 | 0.564 | 19.472 | 0.506 | 6.817 | 0.721 | -6.856 | 0.634 | 2.248 | 0.843 | -10.701 | 0.81 |
| 0.1 | -20.887 | 0.415 | 16.367 | 0.678 | -1.843 | 0.943 | -11.93 | 0.538 | 5.41 | 0.724 | 12.882 | 0.83 |
| 0.2 | -16.002 | 0.648 | 34.044 | 0.528 | 6.382 | 0.856 | -12.215 | 0.645 | 0.219 | 0.992 | -12.428 | 0.879 |
| 0.5 | -20.493 | 0.704 | 49.481 | 0.552 | 5.893 | 0.913 | -22.849 | 0.576 | 2.459 | 0.939 | -14.491 | 0.909 |
| 1 | -31.487 | 0.654 | 46.742 | 0.666 | 5.893 | 0.933 | -32.108 | 0.546 | 2.006 | 0.962 | 8.954 | 0.957 |
| MDD status | -5.51e-04 | 0.634 | -0.002 | 0.226 | 1.48e-05 | 0.99 | -7.14e-04 | 0.422 | 7.97e-04 | 0.249 | 0.003 | 0.333 |

###### Supplementary Table 4. Genomic inflation factor for discovery, adult replication (LBC 1921 + LBC 1936 + ALSPAC adults) and adolescent replication (ALSPAC adolescents) MWAS.

| p threshold | Discovery | Replication (adults) | Replication (adolescent) |
| --- | --- | --- | --- |
| 5e-08 | 0.954 | 0.997 | 1.006 |
| 1e-06 | 0.963 | 0.998 | 0.995 |
| 1e-04 | 0.991 | 0.975 | 1.04 |
| 1e-03 | 0.993 | 0.985 | 1.054 |
| 0.01 | 1.011 | 0.929 | 0.977 |
| 0.05 | 0.997 | 0.958 | 0.946 |
| 0.1 | 0.967 | 0.976 | 0.94 |
| 0.5 | 0.972 | 0.965 | 0.943 |
| 1 | 0.99 | 0.981 | 0.943 |

###### Supplementary Table 5. Top CpG probes associated with PRS at pT = 5e-8. CHR=chromosome, BP=base pair position, SE = standard error, p.adj = Bonferroni-corrected p value, HetChiSq = Chi-square statistics for heterogeneity analysis, and HetPVal = p value for heterogeneity analysis.

| Location | CpG | CHR | BP | Beta | SE | p | p.adj | Direction | HetChiSq | HetPVal |
| --- | --- | --- | --- | --- | --- | --- | --- | --- | --- | --- |
| With MHC region | cg12914966 | 6 | 28830789 | 0.076 | 0.003 | 1.78e-117 | 1.37e-111 | ++ | 148.503 | 3.68e-34 |
|  | cg03270340 | 6 | 28891204 | 0.051 | 0.002 | 1.72e-113 | 1.32e-107 | ++ | 27.397 | 1.66e-07 |
|  | cg00903577 | 6 | 28831109 | 0.058 | 0.003 | 4.64e-113 | 3.57e-107 | ++ | 91.869 | 9.26e-22 |
|  | cg16677399 | 6 | 28830902 | 0.05 | 0.002 | 2.28e-108 | 1.75e-102 | ++ | 210.005 | 1.37e-47 |
|  | cg14345882 | 6 | 26364793 | -0.05 | 0.002 | 4.61e-105 | 3.55e-99 | -- | 157.802 | 3.42e-36 |
|  | cg17849569 | 6 | 28058911 | -0.054 | 0.003 | 1.24e-94 | 9.54e-89 | -- | 72.777 | 1.45e-17 |
|  | cg06608359 | 6 | 28831300 | 0.057 | 0.003 | 6.78e-93 | 5.22e-87 | ++ | 25.551 | 4.31e-07 |
|  | cg12740337 | 6 | 28058973 | -0.045 | 0.002 | 9.95e-90 | 7.66e-84 | -- | 99.663 | 1.81e-23 |
|  | cg27543291 | 6 | 26367644 | -0.053 | 0.003 | 1.59e-89 | 1.22e-83 | -- | 157.825 | 3.38e-36 |
|  | cg10046620 | 6 | 27775042 | -0.041 | 0.002 | 3.61e-89 | 2.78e-83 | -- | 69.556 | 7.43e-17 |
| Without MHC region | cg21600908 | 12 | 133085885 | 0.209 | 0.011 | 6.17e-76 | 4.74e-70 | ?+ | <1e-324 | 1 |
|  | cg05328885 | 11 | 30943623 | 0.035 | 0.004 | 1.28e-20 | 9.85e-15 | ++ | 31.342 | 2.16e-08 |
|  | cg05590274 | 11 | 113262625 | 0.014 | 0.002 | 2.84e-15 | 2.19e-09 | ++ | 11.658 | 6.39e-04 |
|  | cg17862947 | 12 | 133086926 | 0.008 | 1.00e-03 | 3.32e-14 | 2.55e-08 | ++ | 253.185 | 5.25e-57 |
|  | cg09256413 | 1 | 72566690 | -0.011 | 0.002 | 1.52e-13 | 1.17e-07 | -- | 6.382 | 0.012 |
|  | cg06941483 | 11 | 30874744 | 0.014 | 0.002 | 4.08e-12 | 3.14e-06 | ++ | 3.194 | 0.074 |
|  | cg06276712 | 7 | 12107011 | -0.01 | 0.002 | 5.80e-12 | 4.46e-06 | -- | 18.124 | 2.07e-05 |
|  | cg26200795 | 18 | 52895482 | 0.015 | 0.002 | 6.24e-12 | 4.80e-06 | ++ | 2.434 | 0.119 |
|  | cg06627827 | 1 | 72898668 | -0.014 | 0.002 | 1.41e-11 | 1.09e-05 | -- | 1.035 | 0.309 |
|  | cg14159747 | 11 | 113255604 | -0.015 | 0.002 | 5.14e-11 | 3.95e-05 | -- | 14.743 | 1.23e-04 |

###### Supplementary Table 6. Descriptive statistics for CpG locations.

| Relation to Island | Sig CpGs (% in Sig CpGs) | Non-sig CpGs (% in non-sig CpGs) | X squared | df | p |
| --- | --- | --- | --- | --- | --- |
| Island | 0.169 | 0.189 | 1.42 | 1 | 0.233 |
| N_Shelf | 0.045 | 0.036 | 1.165 | 1 | 0.28 |
| N_Shore | 0.159 | 0.097 | 24.727 | 1 | 6.61e-07 |
| OpenSea | 0.479 | 0.562 | 16.184 | 1 | 5.75e-05 |
| S_Shelf | 0.025 | 0.033 | 1.027 | 1 | 0.311 |
| S_Shore | 0.124 | 0.083 | 12.285 | 1 | 4.57e-04 |

###### Supplementary Table 7. Results for discovery MR of DNAm to depression. N_SNP_ = Number of genetic instruments included in the analysis. IVW = inverse‐variance weighted, WM = weighted median, p.adj = FDR-corrected p value, p_Egger intercept_ = p value for Egger intercept evaluation, Q = Q statistics for heterogeneity, p_Q_ = p value for Q statistics.

|  | | IVW | | WM | | MR Egger | | QC measures | | | |
| --- | --- | --- | --- | --- | --- | --- | --- | --- | --- | --- | --- |
| Exposure | N_SNP_ | Beta | p.adj | Beta | p.adj | Beta | p.adj | Egger intercept | p_Egger intercept_ | Q | p_Q_ |
| cg05590274 | 6 | 0.026 | 0.2 | 0.042 | 2.79e-04 | -0.027 | 0.704 | 0.016 | 0.372 | 36.44 | 7.77e-07 |
| cg07519229 | 9 | -0.03 | 1.10e-08 | -0.027 | 4.68e-07 | -0.026 | 0.152 | -0.002 | 0.733 | 15.02 | 0.059 |
| cg08344181 | 18 | 0.045 | 2.99e-47 | 0.048 | 7.57e-48 | 0.057 | 1.23e-06 | -0.008 | 0.049 | 29.86 | 0.027 |
| cg10154826 | 7 | 0.073 | 0.039 | 0.038 | 4.66e-06 | -0.046 | 0.566 | 0.04 | 0.045 | 147.46 | 2.66e-29 |
| cg13813247 | 9 | -0.019 | 0.004 | -0.018 | 1.65e-05 | -0.024 | 0.152 | 0.003 | 0.592 | 27.08 | 6.85e-04 |
| cg14159747 | 8 | -0.029 | 3.75e-05 | -0.034 | 1.39e-07 | -0.034 | 0.152 | 0.002 | 0.748 | 13.32 | 0.065 |
| cg17841099 | 11 | -0.103 | 3.60e-120 | -0.107 | 4.60e-81 | -0.122 | 0.044 | 0.008 | 0.615 | 4.58 | 0.917 |
| cg17862947 | 14 | 0.042 | 8.24e-58 | 0.043 | 4.35e-48 | 0.052 | 5.99e-06 | -0.008 | 0.066 | 18.76 | 0.131 |
| cg17925084 | 6 | 0.037 | 2.29e-04 | 0.04 | 1.10e-06 | 0.055 | 0.203 | -0.006 | 0.546 | 12.77 | 0.026 |
| cg18329187 | 8 | 0.003 | 0.774 | -0.005 | 0.341 | -0.054 | 0.063 | 0.037 | 0.012 | 32.49 | 3.30e-05 |
| cg19624444 | 7 | 0.026 | 5.53e-05 | 0.025 | 2.82e-09 | 0.028 | 0.127 | -0.001 | 0.797 | 16.23 | 0.013 |
| cg19800032 | 5 | 0.046 | 2.26e-04 | 0.048 | 1.97e-08 | 0.011 | 0.739 | 0.011 | 0.315 | 12.69 | 0.013 |
| cg20788561 | 5 | -0.018 | 0.058 | -0.02 | 0.005 | -0.011 | 0.665 | -0.004 | 0.657 | 18.38 | 0.001 |
| cg23275840 | 5 | 0.04 | 1.02e-05 | 0.038 | 2.62e-11 | 0.017 | 0.595 | 0.009 | 0.291 | 13.47 | 0.009 |

###### Supplementary Table 8. Results for replication MR of DNAm to depression. N_SNP_ = Number of genetic instruments included in the analysis. IVW = inverse‐variance weighted, WM = weighted median, p.adj = FDR-corrected p value, p_Egger intercept_ = p value for Egger intercept evaluation, Q = Q statistics for heterogeneity, p_Q_ = p value for Q statistics.

|  | | IVW | | WM | | MR Egger | | QC measures | | | |
| --- | --- | --- | --- | --- | --- | --- | --- | --- | --- | --- | --- |
| Exposure | N_SNP_ | Beta | p.adj | Beta | p.adj | Beta | p.adj | Egger intercept | p_Egger intercept_ | Q | p_Q_ |
| cg05590274 | 7 | 0.166 | 0.016 | 0.153 | 9.59e-05 | 0.047 | 0.75 | 0.01 | 0.341 | 31.23 | 2.29e-05 |
| cg07519229 | 15 | -0.134 | 1.83e-13 | -0.129 | 2.39e-08 | -0.185 | 0.002 | 0.004 | 0.145 | 18.47 | 0.186 |
| cg08344181 | 18 | 0.692 | 1.05e-16 | 0.86 | 4.62e-61 | 0.609 | 0.042 | 0.004 | 0.725 | 100.74 | 6.49e-14 |
| cg10154826 | 27 | 0.228 | 1.19e-09 | 0.154 | 5.51e-09 | -0.039 | 0.75 | 0.031 | 0.03 | 202.31 | 3.18e-29 |
| cg13813247 | 19 | -0.126 | 2.70e-23 | -0.128 | 2.02e-17 | -0.12 | 0.004 | -7.88e-04 | 0.808 | 16.1 | 0.585 |
| cg14159747 | 8 | -0.066 | 0.102 | -0.058 | 0.001 | -0.065 | 0.562 | -1.84e-04 | 0.983 | 37.68 | 3.48e-06 |
| cg17841099 | 11 | -1.022 | 4.21e-119 | -1.059 | 6.48e-62 | -1.545 | 0.005 | 0.022 | 0.164 | 5.98 | 0.817 |
| cg17862947 | 11 | 0.963 | 3.31e-105 | 0.942 | 2.28e-57 | 0.748 | 0.003 | 0.01 | 0.166 | 5.75 | 0.836 |
| cg17925084 | 8 | 0.211 | 1.69e-11 | 0.171 | 3.84e-07 | 0.149 | 0.049 | 0.005 | 0.22 | 9.01 | 0.252 |
| cg18329187 | 17 | 0.05 | 0.036 | 0.022 | 0.37 | -0.019 | 0.697 | 0.012 | 0.039 | 52.57 | 8.86e-06 |
| cg19624444 | 10 | 0.081 | 1.20e-07 | 0.076 | 2.62e-09 | 0.087 | 0.013 | -0.001 | 0.77 | 18.83 | 0.027 |
| cg19800032 | 8 | 0.119 | 0.028 | 0.102 | 0.006 | 0.066 | 0.697 | 0.003 | 0.598 | 26.57 | 3.98e-04 |
| cg20788561 | 11 | -0.13 | 0.002 | -0.16 | 1.23e-04 | -0.109 | 0.327 | -0.002 | 0.777 | 19.97 | 0.03 |
| cg23275840 | 11 | 0.199 | 6.12e-24 | 0.192 | 1.32e-13 | 0.198 | 0.009 | 9.52e-05 | 0.981 | 10.38 | 0.408 |

###### Supplementary Table 9. Results for multivariable MR of DNAm to depression.

| Exposure | N_SNP_ | Beta | SE | p |
| --- | --- | --- | --- | --- |
| cg10154826 | MDD | -0.06 | 0.037 | 0.103 |
| cg14159747 | MDD | 0.024 | 0.03 | 0.421 |
| cg13813247 | MDD | -0.163 | 0.075 | 0.031 |
| cg08344181 | MDD | -0.161 | 0.124 | 0.194 |
| cg19800032 | MDD | 0.039 | 0.046 | 0.396 |
| cg23275840 | MDD | 0.172 | 0.033 | 1.64e-07 |
| cg17925084 | MDD | 0.142 | 0.046 | 0.002 |
| cg07519229 | MDD | 0.004 | 0.078 | 0.96 |
| cg20788561 | MDD | 0.074 | 0.05 | 0.138 |
| cg17841099 | MDD | 0.211 | 0.443 | 0.633 |
| cg17862947 | MDD | 1.024 | 0.414 | 0.013 |
| cg19624444 | MDD | -0.077 | 0.018 | 1.87e-05 |

###### Supplementary Table 10. Results for MR of liability of depression to DNAm. N_SNP_ = Number of genetic instruments included in the analysis. IVW = inverse‐variance weighted, WM = weighted median, p.adj = FDR-corrected p value, p_Egger intercept_ = p value for Egger intercept evaluation, Q = Q statistics for heterogeneity, p_Q_ = p value for Q statistics.

|  | | IVW | | WM | | MR Egger | | QC measures | | | |
| --- | --- | --- | --- | --- | --- | --- | --- | --- | --- | --- | --- |
| Outcome | N_SNP_ | Beta | p.adj | Beta | p.adj | Beta | p.adj | Egger intercept | p_Egger intercept_ | Q | p_Q_ |
| cg05590274 | 122 | 0.027 | 0.157 | 0.008 | 0.564 | 0.011 | 0.9 | 4.38e-04 | 0.851 | 486 | 2.91e-45 |
| cg07519229 | 122 | -0.13 | 0.099 | 0.016 | 0.294 | 0.198 | 0.582 | -0.009 | 0.352 | 7239 | <1e-324 |
| cg08344181 | 123 | 0.104 | 3.61e-04 | -9.48e-04 | 0.932 | 1.02 | 1.39e-17 | -0.025 | 1.12e-15 | 1783 | 1.06e-292 |
| cg10154826 | 123 | 0.172 | 0.004 | 0.01 | 0.812 | 1.926 | 3.08e-15 | -0.047 | 7.93e-14 | 598 | 7.67e-64 |
| cg13813247 | 123 | -0.213 | 0.059 | 0.02 | 0.178 | 0.218 | 0.675 | -0.012 | 0.395 | 13353 | <1e-324 |
| cg14159747 | 123 | -0.008 | 0.763 | 0.008 | 0.677 | -0.002 | 0.984 | -1.43e-04 | 0.963 | 521 | 9.68e-51 |
| cg17841099 | 122 | -0.093 | 1.15e-04 | -0.008 | 0.527 | -0.813 | 8.58e-16 | 0.019 | 9.04e-14 | 1108 | 5.17e-159 |
| cg17862947 | 123 | 0.098 | 3.24e-04 | <1e-324 | 1 | 0.964 | 4.52e-18 | -0.023 | 3.92e-16 | 3063 | <1e-324 |
| cg17925084 | 123 | 0.051 | 0.127 | <1e-324 | 1 | 0.059 | 0.7 | -2.18e-04 | 0.957 | 2085 | <1e-324 |
| cg18329187 | 122 | 0.057 | 0.007 | 0.057 | 0.033 | 0.109 | 0.261 | -0.001 | 0.581 | 145 | 0.068 |
| cg19624444 | 122 | 0.266 | 0.091 | 0.004 | 0.876 | 0.333 | 0.647 | -0.002 | 0.925 | 11975 | <1e-324 |
| cg19800032 | 122 | 0.019 | 0.103 | <1e-324 | 1 | 0.077 | 0.15 | -0.002 | 0.265 | 105 | 0.843 |
| cg20788561 | 123 | -0.108 | 0.118 | -0.015 | 0.308 | -0.216 | 0.493 | 0.003 | 0.724 | 5307 | <1e-324 |
| cg23275840 | 122 | 0.147 | 0.053 | -0.004 | 0.777 | -0.187 | 0.592 | 0.009 | 0.327 | 9730 | <1e-324 |

###### Supplementary Data 1. MWAS summary statistics of PRS for depression at p threshold of 5⨉10_-8._ CpG probes associated with PRS for depression at p threshold of 5⨉10_-8_ were included in the table.

Supplementary Data 2. Results for colocalisation analysis between MDD and CpG probes.
